# Multimodal predictions of end stage chronic kidney disease from asymptomatic individuals for discovery of genomic biomarkers

**DOI:** 10.1101/2024.10.15.24315251

**Authors:** Simona Rabinovici-Cohen, Daniel E Platt, Toshiya Iwamori, Itai Guez, Sanjoy Dey, Aritra Bose, Michiharu Kudo, Laura Cosmai, Camillo Porta, Akira Koseki, Pablo Meyer

**Author notes:** Contributed equally (Simona Rabinovici-Cohen, Daniel E Platt, Toshiya Iwamori).

## Abstract

Chronic kidney disease (CKD) is a complex condition where the kidneys are damaged and progressively lose their ability to filter blood, 10% of the world population have the disease that often goes undetected until it is too late for intervention. Using the UK Biobank (UKBB) we constructed a CKD cohort of patients (n=46,986) with genomic, clinical and demographic data available, a subset (n=2,151) having also whole body Magnetic Resonance Imaging (MRI) scans. We used this multimodal cohort to successfully predict, from initially healthy patients, their 5-year outcomes for End-Stage Renal Disease (ESRD, n=210, AUC=0.804 ± 0.03 with 5 fold cross-validation) and the larger cohort for validation to predict time-to ESRD and perform Genome-wide association studies (GWAS). Extracting important clinical, phenotypic and genetic features from the models, we were able to stratify the cohorts based on a novel set of significant previously unreported SNPs related to mitochondria/cell death, kidney development and function. In particular, we show that the risk allele of SNP rs1383063 present in 30% of the population irrespective of ancestry and putatively regulating *MAGI-1*, a gene expressed in the podocyte slit diaphragm, is a strong predictor of ESRD and stratifies male populations of older age.

## Introduction

Chronic kidney disease (CKD) is a condition where the kidneys are damaged and progressively lose their ability to filter blood. It is estimated that 800 millions or 10% of the world population have CKD [1] and 37 millions in the USA alone. It has been one of the leading causes of death, while 90% of adults with CKD and 40% of adults with severe CKD do not know that they already have the disease [2]. CKD is primarily defined in Clinical Practice Guidelines in terms of kidney function [3] and CKD patients progress over multiple CKD stages, often slowly and heterogeneously [4], from mild kidney damage to End-Stage Renal Disease (ESRD) or kidney failure, defined as either the initiation of dialysis or kidney transplant.

Despite its prevalence, CKD goes often undetected and it is necessary to have better understanding of kidney progression and identifying pre-clinical kidney damage that may lead to CKD. Indeed, the underlying risk factors and pathophysiological mechanisms of CKD have not been well defined. This might be due to the fact that many comorbidities are associated with CKD, the main ones being Type 2 diabetes (T2D), hypertension (HT) and congestive heart failure (CHF), but also, because most of the focus has been on predicting ESRD from late stages of CKD [5, 6, 7, 8, 9, 10, 11], when not much can be done to better understand and slow the progress of the disease. Detecting early-on the deterioration of renal function is an important initial task to then be able to define co-morbdities and their effect on reducing disease burden to finally slow the deterioration of renal function.

Substantial kidney damage as determined by loss of nephrons, is a good example of CKD’s complex etiology as it does not necessarily immediately lead to clinically measurable effects [12], with substantial nephron loss appearing simply through aging [13]. This is due to adaptive responses from remaining nephrons compensating for the missing ones [14], but this also tends to set the stage for more nephron loss [15, 16, 17]. In such cases, imaging may be useful as some larger scale correlations in ultrasound and magnetic resonance imaging (MRI) have been noted [18], and injection of specialized contrast agents with MRI tuning make it sensitive to glomerular microstructures. However, substantial work is still needed to address scan time and contrast agent toxicity for clinical use [19, 20]. In addition, according to a position article which is based on research conducted in the last decade [19], these images can be used to measure volumetric data on the kidney which is another important indicator for CKD progression.

Conversely, rather than using direct markers of specific glomerular and nephron kidney injury, later stages of CKD are defined based on lower levels of creatinine-based estimated glomerular filtration rate (eGFR below 60 ml min 1.73m^™2^) hence capturing an heterogeneous set of kidney disorders. Genomic studies have used eGFR as a trait for finding common variants for kidney disease, seeking possible underlying molecular-level etiology. Such genome-wide association studies (GWAS) have been successful in explaining up to 20% of an estimated 54% heritability in this CKD-associated trait [21] and have helped establish genome-wide polygenic scores (PRS) across ancestries for discriminating moderate-to-advanced CKD from population controls [22]. There is therefore an opportunity to explore the relationship between larger scale image features, clinical measures of kidney function, and genetic analyses for the transition of early CKD to ESRD.

In this study, we applied a multimodal approach to predict, from early stages of the disease, progression of CKD to ESRD. The approach entails using demographic data, clinical data from Electronic Health Records (EHR), single nucleotide polymorphisms (SNPs) and whole body MRI imaging data from UK Biobank (UKBB), a large-scale biomedical database and research resource with half a million UK participants. While a number of studies have sought to identify CKD using Artificial Intelligence (AI) to predict disease, little has been done to predict progression from early-on stages to ESRD, using whole body MRI scans [23], although the possibility of using AI to identify predictive features in image data has been showing some success [24, 25, 26] and has been explored for CKD looking into multiple modalities that inform structural changes and vascular function [27, 28]. Specifically, we sought to identify whether factors influencing the progression could be detected in the integration of genomic data with imaging and clinical data. Such information may aid in early planning of therapy and identification of patients requiring more aggressive treatment and testing, as well as distinguish which genetic variants may tie different aspects of imaging and disease to kidney failure mechanisms and processes. It would also help alleviate the fact that despite superior performance in clinical decision support using multiple data types, a major drawback for widespread adoption of AI models has been the lack of well-defined methods for interpreting such models [29]. To our knowledge this is the first successful study applying a multimodal approach to predict advanced stages of CKD and dialysis from early stages, or even before the disease has manifested. Our approach also led to discovering a new set of genes associated with CKD progression, in particular, to features extracted from radiomic analysis of the kidney and a variant putatively influencing the expression of *MAGI-1* and able to differentiate slow from fast progressors to ESRD.

## Results

According to the most recent update [30], CKD is defined as abnormalities of kidney structure or function, present for a minimum of 3 months, with implications for health; furthermore, CKD is classified based on cause, GFR category i.e. severity of kidney impairment, and severity of albuminuria, the so-called CGA classification. Both the estimation of GFR, as well as the exact definition of the chronicity criterion mentioned in the KIDGO definition, together with physiological and pathological fluctuations of kidney function, represent huge pitfalls in the reliability of the above definition, not to take into account agerelated differences [31, 32], as well as the widely distributed so-called “social determinants of health” [33]. Furthermore, since that definition ultimately may encompass also physiological and para-physiological conditions, one risk at a global level is to devote unnecessary resources to subjects (and not patients) not really needing them. Therefore, an innovative approach to accurately predict, from early stages of CKD, and even before its diagnosis, ESRD in the general population would represent a real game-changer from a medical, as well as a socio-economic, point of view. The overall design and selection of the CKD cohort and definition of the 5-year threshold classification task are presented in Figure 1 and Methods. Briefly, 49,744 patients from UKBB were found to have been diagnosed with CKD (see Methods and [34] for cohort definition), the average age of CKD patients was 67.7 years and the progress to ESRD for the 210 cases present from the date of CKD 1&2 diagnosis was about 70.5 months. We note the cohort average age is slightly older than the UKBB set, suggesting that older patients have had more time for CKD to emerge and be clinically diagnosed. In order to implement multimodal models, this initial CKD cohort was reduced to the 2151 patients for which genomic and MRI information was available, the latter one being the limiting datatype (see Fig.1a). Given the disease progression in this cohort, we decided that a relevant classification task would consist on predicting whether a patient currently, or diagnosed in the near future with early stages of CKD, would progress to ESRD in a 5 year window. The start of the 5 year window, i.e the index date, is defined as the time that an MRI scan was first taken. Notably, when the first MRI scan was taken, none of the patients selected have a CKD 3 & 4 diagnosis and only 188 have a CKD 1 & 2 diagnosis (see Fig.1b).

**Fig. 1:**
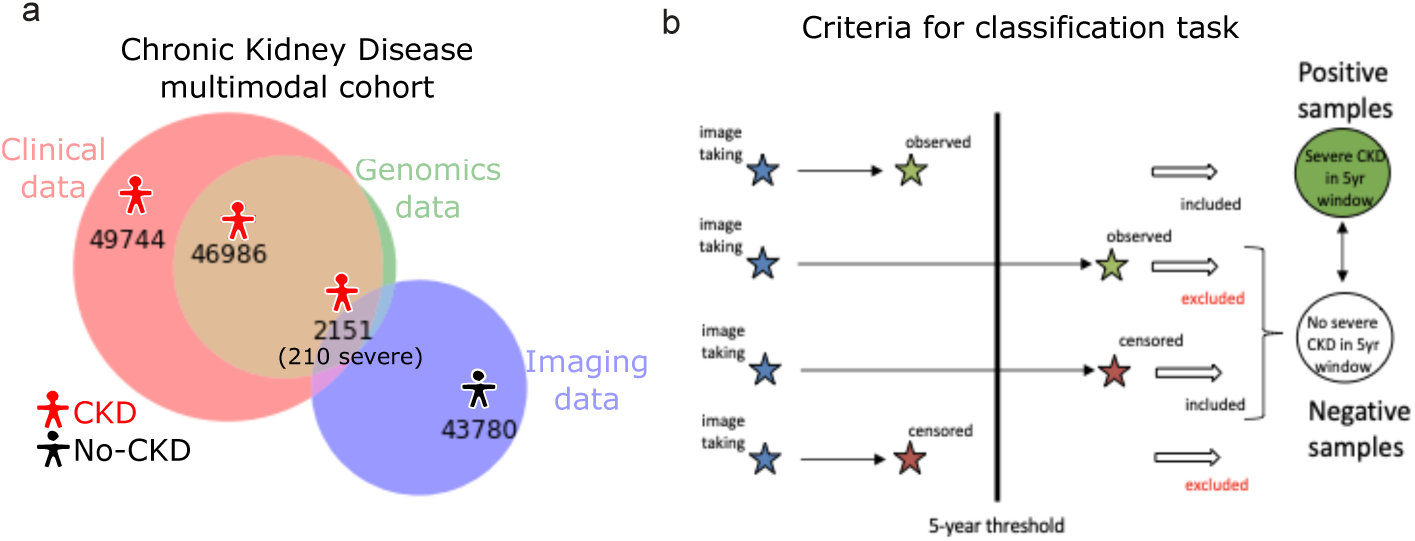
Prediction of End-Stage Renal Disease using a multimodal cohort. **a.** Chronic Kidney Disease (CKD) multimodal cohort definition based on the intersection of patients diagnosed with CKD (49,744), with genomic data available (46,986) and MRI scans (2,151). Out of those patients, 210 reached End-Stage renal disease (ESRD). **b.** Definition of the classification task for progression from early stages of CKD to ESRD. The index date, i.e start time for counting the 5 year window, was set as the first record of an MRI (blue stars). If a patient was diagnosed with ESRD within that window then it was counted as a positive sample (top green star), if diagnosis was done after 5 years (bottom green star) then patients were excluded from the analysis (9 patients with very diverse intervals). Patients censored, i.e not having more records, before the 5 year window were excluded from the analysis (bottom red star) but patients that did not have an ESRD diagnosis within 5 years were counted as negative samples (top red star).

In order to build a multimodal prediction for the 5 year ESRD classification task we implemented 3 types of models, Logistic Regression, Random Forest classifier and XGBoost classifier on features derived from the 4 types of data, demographic, Clinical Classifications Software (CCS) codes, MRI and genomic features. The demographic and clinical features were directly implemented, but the genomic data features were extracted performing a Genome Wide Association Study (see Methods). Likewise, the MRI data was used to extract features from the implementation of three different pipelines of analysis, first the extraction of radiomic features, second a Convolutional Neural Network (CNN) and third a Vision Transformer (ViT) (see Fig.2a). The summary of the results of a 5-fold cross-validation scheme can be seen in Figure 2b and the complete results in Supplementary Figures S1-8. Briefly, age and gender extracted from demographic data were able to predict ESRD 5-year outcome with an AUC of 0.703, radiomics had the best prediction with an AUROC of 0.743 while the other imaging schemes ViT (AUROC=0.657), CNN (AUROC=0.605) and clinical data (AUROC=0.640) had similar performance (Fig.2b). Notably an ensemble method using a voting scheme to integrate all approaches obtained the highest AUC of 0.804*±*0.03 (Fig.2b). Although the GWAS analysis was able to extract 215 significant SNPs associated with CKD, their inclusion as features in the 5 Year-ESRD classification task or using them to calculate a Polygenic Risk Score did not bring any improvement to the performance (AUC=0.54, see Methods). In order to better understand the multimodal predictions, we performed a Grad-CAM analysis [35] for the results of the CNN pipeline and SHAP analysis [36] to rank feature importance for the radiomics features, clinical and demographic data. Interestingly, the attention of the CNN pipeline was mainly concentrated on kidneys and heart (Fig.2c) and clinical terms related to these two organs also appeared as the most important CCS codes in the clinical predictions (see Supplementary Fig. S4b). The SHAP analysis shows that age of diagnosis and sex are very important features for prediction of the disease outcome (see Fig.2d top), while for the radiomics features it shows that the top five Shapley numbers were Energy [37] and Total Energy [38] from the first order statistics, from the Gray Level Size Zone Matrix (GLSZM) Features the Zone Entropy [39], from the Gray Level Dependence Matrix (GLDM) Features the Dependence Non Uniformity [40] and Inverse Difference Moment Normalized (IDMN) [41] (see Fig.2d bottom).Hence, we can interpret these results as having a smaller kidney volume accounted as Energy and overall low image heterogeneity were features strongly predictive of ESRD (see Fig.2d top).

**Fig. 2:**
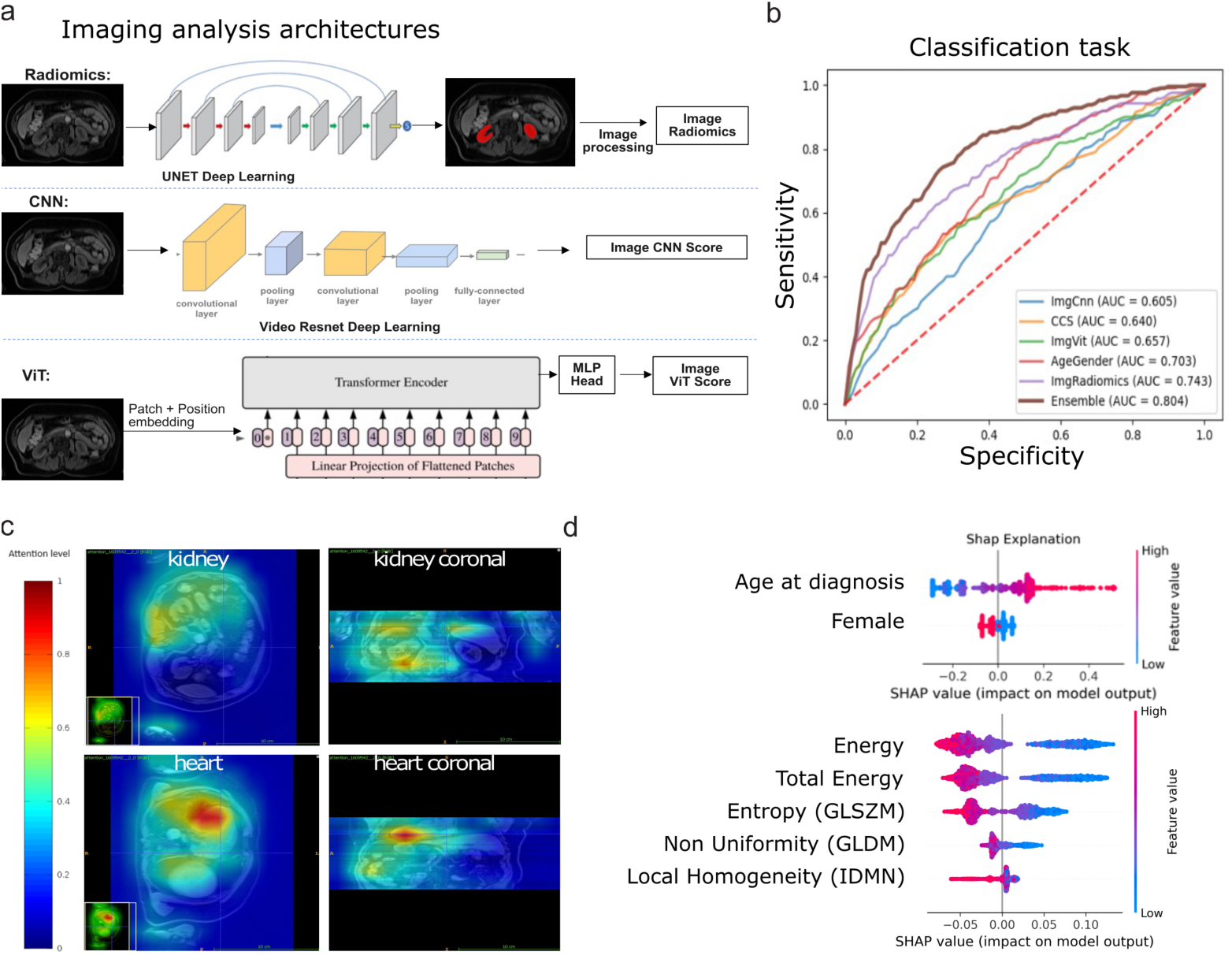
Multimodal prediction of end-stage renal disease from early CKD. **a.** Three types of analysis pipelines for analysis MRI scans, *top* Radiomics *middle* Convolutional Neural Network (CNN), *bottom* Vision Transformer (ViT). See Figures S1-S8 for results details. **b.** AUROC for the 5 year-ESRD classification task with 5-fold cross-validation using each of the data modalities, CNN in blue, Clinical in orange, ViT in green, demographic in red, Radiomics in violet and Ensemble prediction in brown. Genomic is not plotted as AUROC=0.54. **c.** Attention heatmap for the CNN shows kidney and heart being prominent. **d.** SHAP analysis for *top* demographic data and *bottom* Radiomics. Y axis represents different features, heatmap is feature importance for ESRD outcome and X axis is feature value. Energy is a measure of voxel values; Gray Level Size Zone Matrix (GLSZM) Entropy measures heterogeneity in an image; A lower value of Gray Level Dependence Matrix (GLDM) non-uniformity correlates with a greater similarity in intensity values; Gray-level co-occurrence matrix (GLCM) inverse difference moment normalized (IDMN) is a measure of the local homogeneity of an image. The first four features were acquired in water, the last one in fatty tissue. See Tables S11-S12 for Radiomics details.

The analysis of the results for the multimodal predictions for ESRD reveal that radiomics has the largest predictive power and the 215 SNP features extracted from the genomic data are the weakest predictors, also reflected in Odds-Ratio (OR) close to 1 when applying logistic regression for the larger genomic cohort of 46,986 patients (see Fig.1 & Supplementary Fig.9-14). However, these relevant SNPs were extracted for CKD as our cohort only had 210 patients with ESRD an insufficient number for statistically significant GWAS analysis (see Tables S1-S10 for GWAS analysis). To overcome this, we decided to take advantage of our multimodal approach. Indeed, primary setup for GWAS is to compare two groups of subjects against differences in traits. Often, the size of the effect, although significant, tends to be very small due to several factors such as rare variants, complex relationships among SNPs such as epistatic effects, and heterogeneity of the trait. Non-genetic variables such as clinical data, laboratory measurements including eGFR and chemical entities or imaging of body variables are traits that can be used for GWAS. To address trait heterogeneity in ESRD as a possible cause for the low predictive power, we took advantage of our multimodal data approach and the fact that radiomic features had high predictive power to stratify the target population based on the presence of predictive features (see Fig.3a). We hoped that performing a GWAS analysis on this stratified population would allow the discovery of SNP variants associated to ESRD.

Each of the top five SHAP features from the radiomics model were used to stratify the cohort using a binary cut at the mean of the feature value to divide it in two populations (see Fig.2d top). Then, for each of these features, the segmented cohort was used to perform an extraction of SNPs by GWAS, who where then mapped to genes, and Gene Ontology (GO) terms associated with CKD were identified (see Table 1). Gene Ontology enrichment analysis allows the discovery of SNPs focal associations with certain structural components given by the non-genetic data and also helps to overcome the false negative barrier by asking whether the observed association is higher than would be expected given random sampling. Figure 3b & c shows the results of mapping energy and nonuniformity imaging SHAP associated SNPs to “kidney” or “renal” associated GO terms, the only kidney-associated terms that showed statistically significant results and larger OR, after running the analysis with a limited set of 2,151 image samples. Indeed, microstructure features like glomerulus and nephron show weak associations compared to larger scale features such as kidney and renal that the MRI scans are able to detect. However, GO terms associated with kidney or adrenal gland development, filtration and homeostasis, and kidney morphology were present in the genes associated to the relevant SNPs for Energy and Non-Uniformity (see Table 1 and SNPs list in Table S10).

**Fig. 3:**
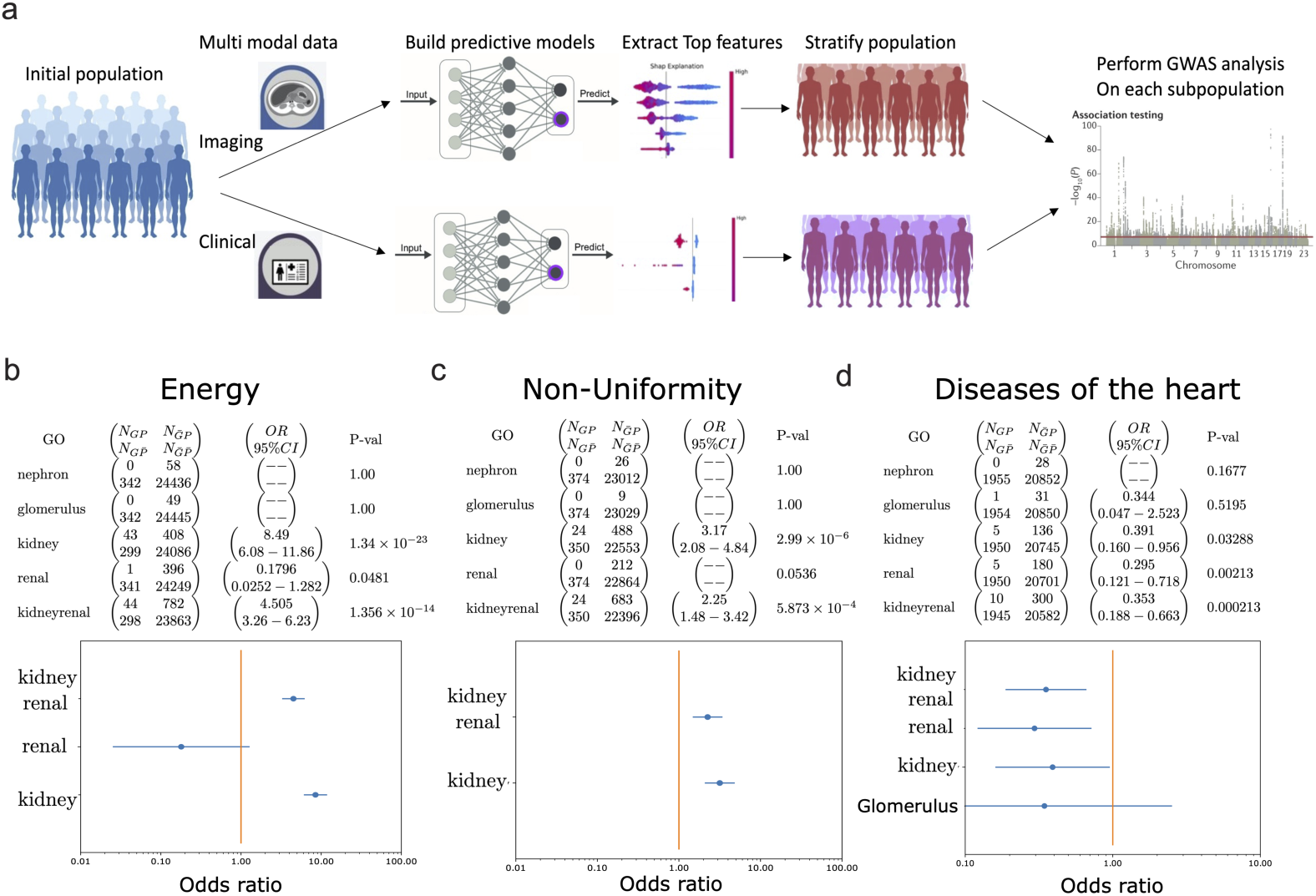
Extraction of relevant SNPs from traits predictive of end-stages of renal disease. **a.** We first constructed unimodal models with *imaging* and *clinical data*. A ranking of important features is then performed for each model using SHAP *middle left panel*. Each of the top features is then used as a trait and a binary stratification of the population is implemented using the feature’s mean as a cutoff and where each patient including the feature is part of the group associated with the trait *middle right panel*. A GWAS analysis is implemented on the specific subset of the population where the top feature is present *right panel*. Results of the Gene set enrichment analysis relative to kidney-related terms for the Energy **b.** and Non-Uniformity **c.** imaging features and **d.** clinical features related to diseases of the heart, mark significant imaging results for most of the GO terms, selecting p-values of 0.001 yielded significant kidney, renal, and kidney+renal GO terms. Shown are the confusion matrix, odds ratio and 95% confidence interval, and Fisher exact test p-value. The Odds Ratio analysis for significant features are shown below and are undefined where *N_GP_* = 0.

**Table 1:** Energy or Non-uniformity SHAP associated SNPs with kidney or renal associated GO-terms.

| Symbol | Count | GO_term |
| --- | --- | --- |
| CASP9 | 81 | kidney development |
| GLI3 | 16 | cell differentiation involved in kidney development |
| CLCNKB | 13 | renal absorption/ sodium ion absorption |
| PPP3CA | 11 | cell proliferation in kidney morphogenesis/renal filtration |
| INSR | 3 | adrenal gland development |
| SALL1 | 3 | kidney/ kidney epithelium/adrenal gland development |
| AQP3 | 2 | renal water homeostasis/absorption |
| ARMC5 | 1 | adrenal cortex development |

| Symbol | Count | GO_term |
| --- | --- | --- |
| CASP9 | 54 | kidney development |
| C1GALT1 | 10 | kidney development |
| ALDH1A2 | 8 | kidney development |
| FREM2 | 3 | kidney development |
| LRRK2 | 1 | regulation of kidney size |
| UPK3A | 1 | kidney development |

Although the stratification approach of the imaging cohort showed novel significant genes related to ESRD, the GWAS analysis is still limited by the only 2151 patients with available MRI data. In order to expand the cohort to obtain a higher statistical power for the GWAS analysis and find SNPs associated to predictive features, we took advantage of the larger CKD population with clinical data and composed of 49,774 patients (see Fig.1). Given that the clinical data did not perform highly on the ESRD-5 year outcome prediction task (see Fig.2b), we decided to implement the time to event model RankSVX [42] that uses a reduced set of clinical features to allow for cohort stratification and interpretable predictions [34]. The time to event task consisted on predicting ESRD onset from stages 1 & 2 of CKD (see Methods). The top features of the predictions, as determined by SHAP analysis, consisted of “Sex” and the level 3 CCS code “Diseases of the Heart” (see supplementary Figure 9) that was used to stratify and implement the GWAS pipeline described in Figure 3a on the 46,986 subset of patients with genomic information (see results Fig.3d). Although most of the patients are censored, i.e only 210 patients reach ESRD (see Fig.4 a & b), the model was able to perform well when predicting the time it takes to progress from early CKD stages 1 & 2 to ESRD, as shown by the concordance index (c-index) and Mean Absolute error (MAE) (see Fig. 4c). As shown in our previous publication [34], using higher level 3 CCS codes did not deter the model performance (see Fig. 4c compare CCS level 3 vs. CCS level 4) and helped obtain a less granular set of features given that all CCS 4 level concepts are included in CCS level 3. This allowed to stratify a population of about 10,000 patients around the top feature “Diseases of the Heart” to implement the GWAS analysis pipeline described in Figure 3a. Genes with significant SNP associations to “Disease of the Heart” SHAP scores with kidney and Cardiovascular disease (CVD) GO-terms are listed in Table 2. They show relatively weak p-values for both terms, yet even at relaxed p-value levels, the GO-terms (Table 2 and Figure 3d) identify consistent structural features relevant to CKD and CVD. Curiously, the GO-terms are anti-enriched that is the Odds Ratio are lower than would be expected by chance when choosing random genes and so the genes containing SNPs associated with the “Disease of the Heart” CCS feature (Figure 3d) are an under-representation of the subset of the CVD and CKD related GO-terms.

**Fig. 4:**
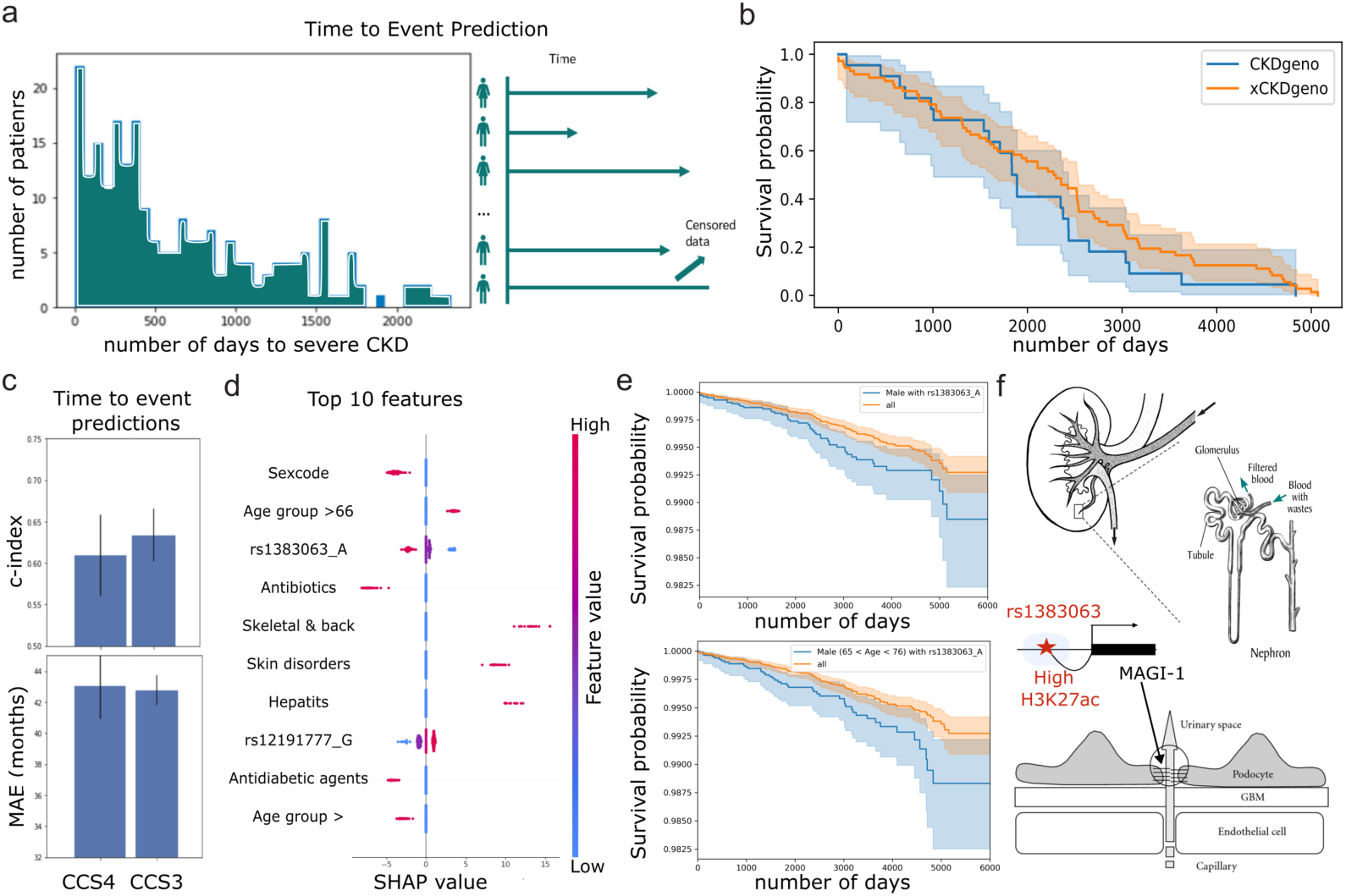
Time to event prediction of End-Stage Chronic Kidney Disease using clinical and genomic features. **a.** Histogram of the number of days elapsed from early CKD diagnosis (stage 1 or 2) to ESRD for the 210 patients in UKBB cohort. Right diagram shows the Time to Event prediction task including censored data, i.e patients diagnosed with early CKD but that have not progressed yet to ESRD. **b.** Survival curve for 94 patients from **a.** conditioned on whether they have any of the SNPs (23 patients) indicated as CKDgeno in blue curve or with none (71 patients) indicated as the orange curve xCKDgeno. Differences are not statistically significant. **c.** Performance of RankSVX model using Clinical Classifications Software (CCS) codes level 3 or 4 as features is shown using c-index and Mean Absolute Error (MAE). **d.** Top 10 features of RankSVX model using CCS3 and genomic features extracted by SHAP. Red dots represent the feature value and are an indicator of having the indicated disease (CCS205 Spondylosis, CCS200 Other skin disorders, CCS6 Hepatitis), a relevant SNP variant (rs1383063 A or rs12191777 G), being prescribed with an associated drug therapeutic class (Antibiotics erythromicin and macrolides or Antidiabetic agents) or a demographic variable (Age higher than, Sex-red indicates male). Negative SHAP values (x-axis) indicate prediction of a higher risk of ESRD. **e.** Survival curves for time to ESRD in blue, male having rs1383063 A *Top* and older than 65 years *bottom*. Orange represents the rest of the cohort. Differences are statistically significant *Top* Pval*<* 0.0139, *bottom* Pval*<* 0.0085. **f**. SNP rs1383063 sits 50kb upstream of the gene MAGI-1 in a potential enhancer element enhD E2210115 that has been shown to be acetylated in H3K27 see *middle*. MAGI-1 is expressed in the podocyte slit diaphragm shown in *bottom* part of the kidney glomeruli shown on *top*. GBM stands for Glomerulus 15 Basement Membrane.

**Table 2:** ”Diseases of the heart” SHAP associated SNPs with CVD (italicized), kidney, or renal associated GO-terms for GWAS SNP p-values = 0.001.

| Symbol | Count | GO_term |
| --- | --- | --- |
| MME | 4 | kidney development/*cardiolipin binding* |
| ARMC5 | 3 | adrenal cortex development |
| BCL2 | 1 | renal system process |
| ADORA1 | 1 | negative regulation of renal sodium excretion/*heart contraction* |
| WDPCP | 1 | kidney development |

That would suggest that the specific issues that the clinical analysis identified for cardiovascular problems are most significantly tied to a small and specific subset of genetic kidney problems probably defined by genes *MME, ARMC5, BCL2, ADORA1* and *WDPCP* (see SNPs list in Table S10). Interestingly, both MME and BCL2 activity are cardiolipin-dependent, a lipid mainly found in mitochondria and heavily enriched in cardiomyocytes [43]. These add to the list of mitochondria-localized proteins CASP9 (Table 1) and ACSF3 (Table S8), part of the mitochondrial fatty acid synthesis (mtFAS), a highly conserved pathway essential for mitochondrial biogenesis [44]. Overall, these genes illustrate how SNPs related to genes performing mitochondria-related functions such as metabolism and apoptosis have been found to be over-represented in our analysis (see Table S7) and might underlie that regulation of cell death in the kidney is an important characteristic of CKD severity.

Given the success of the survival analysis for extracting genes relative to specific subset of kidney problems, we decided to test the predictive power of the 215 SNPs associated with CKD (see Tables S7 & S8) on this time to event task. Indeed, although after censoring, the number of subjects in the 5 year ESRD prediction cohort who actually progressed to ESRD is small and these genetic features yield relatively low power, we observe a systematically higher rate of conversion to ESRD for subjects carrying any of the 215 SNPs as shown in the Kaplan-Meier curves (see Fig.4b). Hence, we combined the 215 CKD-associated SNPs with the clinical features from the cohort of 46,986 patients with CKD and trained a RankSVX model for the time to ESRD task. The SHAP analysis for the top predictive features included sex and age, already shown to be important predictors (see Fig.2d and Fig.S17), but also two SNP loci were included, rs1383063 ranked 3rd and rs12191777 ranked 8th (see Fig.4d). Importantly, the top 3 features, being male with age above 65 years and presence of rs1383063 A, could be used to differentiate the outcome of patients in a statistically significant way, as shown by the Kaplan-Meier curve (see Fig.4e and see Supplementary Figure S18). Also, although genes near rs12191777 did not have kidney-related functions and did not reach genome-wide significance ((see Supplementary Figure S10), rs1383063 falls in a cis-Regulatory Elements (cCREs), the distal enhancer E2210115 shown to be acetylated in H3K27, and about 50kb upstream of the kidney-related *MAGI-1* gene and all features fall in the same Topological Associated Domain (see Supplementary Figure S20). In [45] rs1383063 was found to be associated with eGFR/creatinine levels, and in a eQTL but not for *MAGI-1*, however Rap1 pathway, whom MAGI-1 is part of, was reported to be enriched in eGenes from an eQTL study using glomerular and tubulointerstitial samples [46]. Hence there is evidence for rs1383063 SNP being a potential regulator of the expression of *MAGI-1*, whose product is a member of the membrane-associated guanylate kinase homologue (MAGUK) family, participating in the assembly of multiprotein complexes on the inner surface of the plasma membrane at regions of cell-cell contact (see Fig.4f). MAGI-1 protein may play a role as scaffolding protein at cell-cell junctions and in the kidney it has been shown to localize at the podocyte slit diaphragm, a specialized intracellular junction that is universally injured in proteinuric diseases [47, 48]. Precisely, MAGI-1 was found to be differentially expressed in podocytes of CKD vs Control samples in a recent integrated snRNAseq, snATACseq, and scRNAseq study [49]. It has also been shown that diminished MAGI-1 expression in cultured kidney podocytes weakened tight junction integrity, although knock-out mice demonstrated normal glomerular histology, lowering nephrin levels resulted in spontaneous glomerulosclerosis and low levels of MAGI-1 are related to proteinuric states (see Fig.4f bottom) [48, 50]. Interestingly, rs1383063 A was not only present in 30% (Table S8) of UKBB population but was over-represented in different ancestries (see Table 3) even if these minority populations did not play a role in the association of SNPs with CKD (see Supplementary Figure S16).

**Table 3:** Distribution of MAGI-1 regulating alleles in UKBB population.

| **rs1383063-1** | **rs1383063-2** | **# individuals/PP** |
| --- | --- | --- |
| G | A | 149/63882 |
| A | A | 135/41290 |
| G | G | 18/25171 |

| **rs1383063-1** | **rs1383063-2** | **# individuals/PP** |
| --- | --- | --- |
| G | A | 1472/63312 |
| A | A | 705/39967 |
| G | G | 112/25077 |

## Discussion

CKD is a relentless chronic and progressive condition that has been estimated to affect more than 850 million individuals worldwide [51]. Furthermore, epidemiologic studies have shown that CKD has also emerged as a leading cause of global mortality [52, 53], despite some survival improvement recorded in recent years, for end-stage renal disease (ESRD) patients, at least in rich, industrialized, countries [54]. Thus, beyond finding more effective treatments, the early identification of CKD would be of paramount importance; all the more given its global prevalence and although more challenging, it would be necessary to find methods that predict the evolution of CKD into ESRD.

Taking advantage of the UKBB dataset, we were able to build a CKD cohort of more than 2,000 patients to build a multimodal model that is able to effectively use imaging features, in addition to demographic and clinical data, to predict 5-year ESRD outcome with a AUC above 0.8. The radiomic imaging features show that having a smaller kidney volume and overall low image heterogeneity is strongly predictive of ESRD, together with age and sex (see Fig.2d top). Conventionally, nephrologists tend to precisely use kidney length, volume, cortical thickness and echogenicity to evaluate the severity of kidney injury. Very short renal length (e.g., *<* 8 cm), apparent white cortex, and contracted capsule contour, all indicate an irreversible kidney failing process with high specificity but limited sensitivity [3]. The performance of our model is also notable because the vast majority of the patients do not initially have a CKD diagnosis at the time the MRI was performed, or are at very early stages of the disease. Furthermore, by expanding the cohort beyond MRI data to about 50,000 patients with relevant genomic and clinical features, we were able to confirm the results observed in the imaging cohort that age, sex and heart/kidney conditions are the best predictors of the disease outcome (see Figure 4d, Supplementary Figure S4b and S17b). We homed in on a particularly interesting gene *MAGI-1*, regulated by the rs1383063 SNP locus associated with a regulatory distal enhancer and showing a strong predictive effect for males of older age. The fact that *DGL2* also a member of the MAGUK family [55] was part of the top genes of the initial CDK-centered GWAS (Table S8) proves that the survival analysis, unlike the 5-year classification task, was able to distill variant rs1383063 A, with a strong predictive power and present in more than 30% of the population for all ancestries. Overall our study shows that by changing the predictive task from eGFR levels to predicting CKD or ESRD, we obtain an interesting set of new gene candidates associated with CKD and kidney features, such as kidney development and function, as well as mitochondria-related functions of metabolism and apoptosis not reported in previous studies [56, 22]. Also, we think that the phenotype regressions performed against T2D, CHD and HT, helped focus our genetic analysis on CKD itself and hence obtain a very different set of relevant SNPs. Overall, our approach was successful as we are able to train a highly performing multimodal model that is predictive of ESRD from early stages of the disease and show that the MAGUK family of genes *DGL2* & *MAGI-1* are probably good therapeutic targets, given their importance in regulating proteinuria, a common signature of late CKD progression. This important advancement should have clinical and medical impact on the prevention and treatment of chronic kidney disease.

## Data Availability

All data produced are available online at https://www.ukbiobank.ac.uk/

## Acknowledgments

We thank Krzysztof Kiryluk, Guillermo Cecchi and Mayra Furlan-Magaril for their comments while reading the manuscript. This research has been conducted using the UK Biobank Resource under Application Number 95318. Also, this work has been partially funded by the EU Horizon 2020 project CAPABLE #875052.

## Methods

### Code availability

Code is available at: https://github.com/jeriscience/CKDprediction

### Enrollee Selection: Enrollee, GP, and HESIN Records Processing

The UK Biobank (UKBB) recruited around 500,000 people aged between 40-69 years in 2006-2010 from across the UK. With their consent, they provided detailed information about their lifestyle, physical measures and had blood, urine and saliva samples collected and stored for future analysis. UK medical records were consented for inclusion in the UKBB. Data are derived from the UK Biobank’s (UKBB) imputed genomic repository(111,480 subject records), and subjects with imaging data, enrollment intake records, and GP and HESIN records (136,749 which also included all the genotyped data then available). We accepted CKD diagnoses and staging from the UK Biobank rather than eGFR directly since eGFR by itself does not meet clinical practice guidelines: Guidelines specify repeated eGFR with proteinuria testing with persistence over a period of time [3]. A final subset of 2151 CKD patients and 4108 controls corresponding to the intersection of clinical and genomic (68,781 patients, 26,814 controls after removal of duplicated records) and imaging participants (2151 patients) were available for the multimodal analysis components.

### Clinical Overview

We extracted records for CKD stage diagnosis, and for a number of clinically relevant factors, and constructed derived features. Stages were pooled into CKD12 for stages 1 and 2, CKD3, CKD4, and CKD5 (dialysis dependant). First CKD diagnosis did not start at stage 1 for all patients. The earliest date of diagnosis across all states was taken as date of CKD diagnosis. Among these others were diagnosis (Dx) for hypertension (HT), type II diabetes (T2D), congestive heart failure (CHF), whether the Dx for these conditions were applied prior to the diagnosis for chronic kidney failure (CKD) at any stage (preHT, preT2D, preCHF). Sex followed the UK Biobank coding (0 = Female, 1 = Male), age was taken at 2022 derived from UK BB’s date of birth (dob), with a thresholded age (t age) for individuals over 60 (near the mean age of CKD diagnosis), and a centered and scaled age (s age). CKD5 and Dialysis were marked as end stage renal disease ESRD. Codes for Black British (BlackBrit) were identified by UK Biobank codes (4, 2001, 4001, 4002, 4003) and generally South Asian (SouthAsian) UK Biobank codes (3001, 3002, 2003, 3003, 4). See Table S1 for details.

### Adjustment variable selection

We performed logistic regressions predicting diagnosis of chronic kidney disease using pre-CKD hypertension, Type II Diabetes, Congestive Heart Failure, sex, age, Black British status, South Asian status, and age of first CKD diagnosis (Figure S11, Tables S2).

Analysis of population stratification effects are presented here. QQ plots (Figures S9) were prepared first for a shuffed set (all the demographic and clinical data were shuffed in their. columns, Figure S9a). For a random variate *R*, define a cumulative distribution function *F* (*r*) = P(*r/leR*). Then define an r.v. *Q* = *F* (*R*) so that P(*q/leQ*) = P(*r/leR*) = *F* (*r*) = *q*, so *Q* = *F* (*R*) is uniformly distributed. The logistic regression model should uniformly sample the shuffed data, which is in fact seen in the figure. The next test was to establish the baseline without considering population stratification. This included adjustments for age, sex, hypertension diagnosis, type II diabetes diagnosis, and congestive heart failure diagnosis all diagnosed prior to onset of CKD. Ideally, the hope is for stronger single nucleotide polymorphisms with strong p-values that would deviate from the shuffed uniform agreement. This resulting plot (Figure shows much higher associations throughout the range of SNPs than expected by chance with an expansion factor of *lambda* = 2.157, suggesting that there are ancestrally linked deep associations with CKD among subpopulations in the UK Biobank cohort. Population variables employed by the UK biobank (https://biobank.ctsu.ox.ac.uk/crystal/field.cgi?id=21000) is based on UK governmental census and records systems designed to quantify benefits eligibility required by law (https://www.ons.gov.uk/census/census2021dictionary), which may not optimally represent ethnic lineages appropriate for genetic analysis. This question is expanded later in the analysis. Including adjustments for UK Biobank Black British and South Asian categories as listed above, the QQ plot (Figure S9c) still shows significant deviation from the null model, but much of the population based stratification was removed, with the expansion factor now reduced to*, λ* = 1.663. This leaves the difficulty that other identified populations may not reflect clinically relevant heritage classifications suitable for adjustment or stratification. It is worth noting that the impact of Black British and South Asian inclusion was disproportionate to the population sizes represented in the cohort, with Back British showing 243 subjects in the regression, and South Asians showing 2010. Overall, the Manhattan plot for the population adjusted regression (Figure S10) showed relatively high levels of “noise” marking stronger p-values than would be expected by chance, echoing the strong expansion factors. There were relatively few SNPs rising to Bonferroni significance. Given these results, we sought a method that would focus on CKD relevant genes. We therefore chose to filter SNPs according to membership in Gene Ontology records with terms that included nephron, glomerulus, and renal. This reduced the Bonferroni threshold, and focused on any SNVs that were relevant to CKD, whether or not they were impacted by population lineage stratification effects, and offered a measure of relevance for the composite marker by enrichment.

The Age of first CKD diagnosis feature was incompatible with the South Asian feature; logistic regressions converged for CKD, but not for ESRD. CKD and ESRD regressions are displayed in Figures S12a and S12b. Age is a risk to CKD, but not as important for ESRD progression. Both South Asian and Black British are risk factors for CKD, but while being Black British is also a risk for progression to ESRD, being South Asian does not show a similar risk. Being male was a risk for progression to ESRD, but being female was a risk for CKD. The impact of hypertension, type II diabetes, and congestive heart failure were consistent in spite of the other variations. Since we hoped to find relevant Single Nucleotide Polymorphisms (SNPs) that may be ancestrally informative, and the other variables may show relevant interactions with variants, we sought to control only for HT, T2D, and CHF in the GWAS by inclusion as adjustment variables.

We also sought to identify relevant impacts of pharmaceutical therapies to test them as adjustment variables that impact progression. We therefore applied survival analysis (Figure S13), with Kaplan-Meier regression (Figure S13a), and Cox regression (Figure S13b and Table S3), showing the Cox prediction for thiazides. Clinical practice guidelines [57] for diuretic therapies of CKD recommend thiazides early in treatment. The data testing for time-dependent effects included thiazides, and were applied to early CKD stages 1 and 2. The Kaplan-Meier plot shows a tendency for early delay in progression to ESRD. However, the statistical power was not enough to resolve a significant contribution in the Cox regression hazard ratio model (Figure S13). Age of diagnosis significantly increased the hazard ratio, while age significantly protected against disease progression to ESRD.

### Genomic Quality Control and Record Selection

We performed genomic quality control (QC) on the UKBB imputed genotype data of 113,939 samples and 44 million high quality imputed variants (INFO *>* 0.3) using PLINK [58]. We removed missing samples and variants with 2% missing values, minor allele frequency (MAF) *<* 0.05, Hardy-Weinberg Equilibrium (HWE) *<* 10^™6^ and removed samples with gender discrepancy, more than three standard deviations in heterozygosity rates along with closely related individuals (identity by descent) (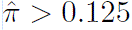). We finally obtained 136,749 samples and 4.44 million variants after QC, from which 46,986 intersected with the 49,744 patients having a CKD diagnosis. 55,896 participants with no CKD history at all and used as controls for GWAS.

### Genomic Analysis

We started by testing Chronic Kidney Disease (CKD) risk covariates, such as Hypertension (HT), congestive heart failure (CHF), type II diabetes mellitus (T2D), race, sex, age, and time of diagnosis, using the logistic regression package from Statsmodels [59], where HT, T2D, and CHF diagnoses prior to CKD diagnoses were taken to exclude diagnoses possibly caused by CKD. We used “lifelines” [60] for Kaplan-Meier and Cox regression analysis to understand the impact of phenotypes and variants on time to ESRD.

Significant predictors of CKD not including age, sex, or race, i.e hypertension, type II diabetes, and congestive heart failure, were included as adjustment variables in the GWAS, computed using PLINK [58]. SNPs with 1 *×* 10^™4^ significance after Benjamini-Hochberg FDR adjustment were retained (see Table S10).

Resulting SNPs were assigned to genes using GeneLocator [61], assigning SNPs within 10kb margins upstream and downstream. We tested Gene Ontology (GO)-terms, using tables acquired through GOATools [62], associating SNPs with GO-terms “kidney,” “nephron,” “glomerulus,” and “renal”. These GO-terms were tested for enrichment predicting CKD. We identified active alleles, and coded these categories if any SNP mapping to the terms was heterozygous or homozygous in the active alleles, and applied logistic regressions to these predicting CKD, and explored time to ESRD using COX regression for these variates. We also explored the GO-terms of the SNPs with the strongest (smallest p-value) associations with CKD, defined a category with heterozygous or homozygous active alleles among assigned GO-terms, with logistic regressions predicting CKD, and COX regressions for time to ESRD. For these regressions sex, age of diagnosis, current age, and time to ESRD were also included.

We also sought to identify genomic features associated with kidney function as defined by the clinical data and structure, using the radiomic imaging analysis identifying kidney disease, that show predictive power in the progression to ESRD. We applied GWAS adjusted by prior hypertension, type II diabetes, and congestive heart failure. SNPs were mapped to genes, and the genes linked to GO-terms as shown in Table 1 as well as Figure 3.

GO enrichment, displayed in Table S4, shows Fisher Exact Test p-values and confusion matrix for gene categories comprised of associated GO terms that contain the “Type” labels in the term as the exposure associating with CKD SNPs with raw p-values *≤* 1 *×* 10^™4^. While these SNPs are far from genomewide significant, they do tend to show highly significant association with kidney function related GO-terms (see Table S10). Interestingly, pooling kidney and renal classifications together (“kidney/renal”) reduced significance.

The false discovery rate was controlled for the resultant SNPs by use of Benjamini and Hochberg’s algorithm. At the 1 *×* 10^™4^ significance level after Benjamini-Hochberg adjustment, 215 SNPs were identified as CKD significant SNPs (see Table S10). However, no SNPs were significant in predicting progression to ESRD in this dataset, probably due to the small numbers of patients in the outcome (n=210). The alleles that were protective vs. deleterious were sorted out, and samples homozygous with deleterious alleles or which were heterozygous were identified as carrying a “CKD genotype” or CKDgeno.

We performed logistic regression (Table S5 and Figure S14) predicting CKD (Table S5a and Figure S14a) and progression to ESRD (Table S5b and Figure S14b) including the same covariates that were included in the phenotypic logistic regressions along with the “CKD genotype.” After inclusion of the other covariates, CKDgeno was actually slightly protective against CKD. It therefore interacts with some of the other covariates. Progression to ESRD showed a trending association with CKDgeno.

Survival analysis regressions [60] were displayed in Table S6 and Figure S15. The Kaplan-Meier regression suggest the impact of CKDgeno on progression is within bounds of the error bars (Figure S15a), which is born out in the pvalues (Table S6) and the error bar plot (Figure S15b). After censoring, the number of subjects in the UKBB who progressed to ESRD had relatively small numbers, yielding relatively low power marked by wide error bars and large p-values. With that caveat, there is a suggestion of a systematically higher rate of conversion to ESRD for CKDgeno carrying subjects (Figure S15c)

In Table S7 and Table S8, are shown the enriched GO terms and the genes associated to the 215 relevant SNPs associated to CKD at *p ≤* 1 *×* 10^™4^ significance (see list of SNPs Table S10). Out of the top genes 20 genes, only 6 have been previously reported. It is only when we lower the significance to the 0.05 level after Benjamini-Hochberg control, that we identified among significant SNPs the following previously published genes [63, 64, 65, 22, 21] *PKD1, PKD2, OGG1, VEGFA, MTHFR, TNF-α, COL4A5, COL4A4, COL4A3, NAT8, SHROOM3, DAB2, WDR37, WDR72, UMOD, TTR, LINC00923, HLA-DQA1, ICAM1, TGFB1, VAV3, DEFA, ITGAM, SLC22A2, CUBN, AFF3, CDCA7-SP3, SCAF8, MYO16-IRS2, RGMA-MCTP2, PLA2R1, APOL1, CYP11B2, AGT, SOD1, SOD2, CAT, GPX1, GPX3, GPX4, IL-1A, IL-4, IL-6, IL-10, ICAM-1*.

### Principal Component Analysis

We performed Principal Component Analysis (PCA) on the QC data after pruning variants for linkage disequilibrium (*r*^2^ *>* 0.25) with using `TeraPCA` [66] and obtained the top fifty principal components (PCs). The results applied to the cohort are shown in Figure S16. The population structure is displayed in Figure S16a. While the prevalence of CKD across the range of genetic variation is well represented (Figure S16b), the pooled CKD geno SNPs are not so evenly represented in minority populations (Figure S16c). We applied logistic regression to identify how the principal components predicted CKD, and to understand whether CKD risk genotypes were biased in its representation across the PC mapped variances (Figure S11 and Table S9). We found that PC0 was mildly predictive of CKD (OR = 1.163, 95%CI: 1.147-1.179, *p* = 5.227 *×* 10^™103^; Table S9a and Figure S11b). The question of whether CKD geno adequately represents minority community genetic variations was tested with another logistic regression (Table S9b and Figure S12). In this, PC0 is strongly under-represented in CKD geno SNPs (OR = 0.602, 95%CI: 0.580-0.626, *p* = 2.832 *×* 10^™147^).

## Image Data Analysis

### UK Biobank MRI data

Subjects invited to neck-to-knee body MRI were recruited by letter from the National Health Service and scanned at three different imaging centres in Great Britain with a Siemens Aera 1.5T device, using a dual-echo Dixon method [67]. The device acquired overlapping images in six stations covering the body from neck to knee within about 6 minutes with TR = 6.69, TE = 2.39/4.77 ms, and flip angle 10deg [68]. The reconstructed, volumetric station images encode voxel-wise intensity values with a separate water, fat, in and opp signal (UK Biobank field 20201). The head, arms, and lower legs extend outside of the field of view and are often distorted near the image borders. The kidneys are typically located in the second and third imaging stations, each of which were acquired in a 17s breathhold with typical dimensions of (224 x 174 x 44) voxels of (2.232 x 2.232 x 4.5) mm.

The MRI stations volumes for all sequences (water, fat, in and opp) were extracted from a downloaded zip file from the UK Biobank servers. The station numbering was determined by sorting the z axis physical coordinates. Volume fusion and interpolation was done by using Langner et al. algorithm [69].

We created three types of models for the imaging data: radiomics image processing model, CNN deep learning model, and a transformer deep learning model. Each model revealed different features of the imaging data. For creating these models we used the FuseMedML open-source framework [70], image analysis methods similar to [71, 72], and a proprietary package planned to be published in the future.

### Radiomics model

*Radiomics* is a quantitative approach to medical imaging. It’s goal is to find associations between qualitative and quantitative information extracted from clinical images and clinical data by using analysis methods from the field of computer vision, information theory and statistics.

For any radiomic approach, it is critical to define the volume of interest (VOI) in a three-dimensional (3D) volume from which the radiomics features will be calculated. Kidney 3D segmentation to be used as VOI to extract radiomics were generated by a pretrained segmentation model (2.5D U-net) from previous research by Langner et al. [69]. The model failed to create segmentation for some patients due to image artefacts in these two stations, such as water-fat swaps, background noise, metal objects, but also non-standard poses, misalignment in the scanner, and corrupted data were excluded after visual inspection of mean intensity projections.

### Radiomics image processing

MRI imaging modalities contain Gaussian and Rician noise and could benefit from de-noising [73]. In addition, medical imaging sequences can be acquired with different protocols, MRI machines and on different sites. As a result, imaging datasets can include non-uniform pixel spacing, signal intensity ranges and so forth. Prior studies have indicated that the robustness of radiomic features is dependent on image processing settings, thus standardisation of the acquired dataset is required. Standard image processing will include interpolation to a isotropic pixel-spacing, intensity range normalization, and discretization [74, 73].

The final image processing is feature extraction, where feature descriptors are computed from the VOI. Zwanenburg et al. [73] proposed dividing radiomic features into a number of feature-families such as morphological features, intensity features, grey level features. Table S11 shows the parameters that were used for radiomics image processing.

All supported feature-families in pyradiomics python package [75] were extracted for each of the four MRI sequences separately.

### Video-resnet CNN and ViT deep learning models

In addition to the kidney segmentation and radiomics feature extraction pipeline, a video-resnet [76] convolutional neural network (CNN) pipeline and a visiontransformer (ViT) [77] pipeline were used to train the MRI images to predict directly the clinical target using the Adam optimizer [78], and the network and hyper-parameters described in Table S12.

### Imaging and clinical ensemble model

In the final stage, we created an ensemble of the three models based on imaging data and two additional models based on clinical data, as kidney clinical data analysis was proved to be significant [79]. The extracted features from each imaging or clinical model were the input to a classical machine learning classifier (logistic regression, random forest, xgboost) to predict our target. When the number of input features was large, we first applied a feature selection method to select the most significant features before applying the classifier. Supplementary figure S1 depicts the ensemble method averaging the scores of all the 5 models (3 imaging models and 2 clinical models).

## EHR Analysis: Time to End-Stage Renal Disease (ESRD)

### Mapping diagnosis features into higher level concepts

Using electronic health records (EHR), patients diagnosed with CKD were selected from the UKBB database, excluding those with pregnancy. These EHR data were, for some patients, encoded as ICD9 while for others were coded using ICD10 codes; therefore, diagnosis codes were normalized by mapping the ICD codes to AHRQ CCS (Agency for Healthcare Research and Quality – Clinical Classifications Software) codes[80]. CCS are composed of diagnostic categories organized in a hierarchical system consisting of four levels for diagnoses^1^. Level 4 has 281 diagnosis codes, level 3 has 134 codes, level 2 has 18 codes, while level 1 has only one code. The CCS codes in our cohort are the leaf nodes in the hierarchy. To overcome the sparsity in the data, we mapped CCS diagnosis codes (the leaf nodes) into higher levels (level 2) [34]. Mapping to level 2 of the CCS ontology helps to interpret the important features driving the progression to ESRD.

### Time-to-ESRD model

In our analysis, the objective of RankSVX[42], a time-to-event model, is to predict the risk and time of ESRD among subjects in stages 1 or 2 of CKD. Unlike other time-to-event models, RankSVX optimizes two functions simultaneously, one to rank subjects based on their risk to event, and the other to predict the actual time to event for non-censored subjects.

For a patient *i*, let us assume that *t_i_* is the duration between the index event (the onset of stage 1 or 2) and the outcome event (ESRD) for non-censored patients or the duration between the index event and the last follow-up visit for censored patients. *x_i_* is the feature vector for patient *i*. RankSVX optimizes the following objective function:

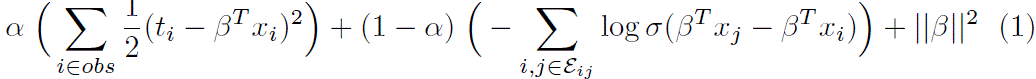

where *β* is the parameter of the linear predictor and *α* is a hyper-parameter to weight each term. The first term optimizes the model to correctly predict the actual time to ESRD for observed patients. The second term optimizes the model to correctly rank the relative risks of two subjects, where *σ* is the sigmoid function and *ε_ij_* represents all pairs of subjects *i, j* where subject *i* observed the event and subject *j* may or may not have observed the event and *t_i_ ≤ t_j_*. The last term is a regularization term to prevent overfitting. More details about the model can be found in a prior paper[42].

### Identifying highly ranked features

After implementing the RankSVX model to predict the time to ESRD, we apply SHAP analysis to identify the top important features [81]. SHAP (SHapley Additive exPlanations) is a method to explain individual predictions, which is based on the game theoretically optimal Shapley values. The goal of SHAP is to explain the prediction of an instance by computing the contribution of each feature to the prediction. The top important features extracted by SHAP are considered as the driving or correlated features to the progression of CKD.

### Stratification and validation

After identifying the top important features (diagnosis and drug codes), we stratify patients based on whether they were assigned with these codes. To model the interactions among features, we also stratified patients based on different combinations of the selected features. In our experiments, we chose the top three features using the SHAP analysis and tested all subgroups of different combination of selected features. Then, we used Kaplan-Meier to assess the correlation of the selected subgroups to the progression of CKD to ESRD.

## Supplementary Tables

**Table S1:**
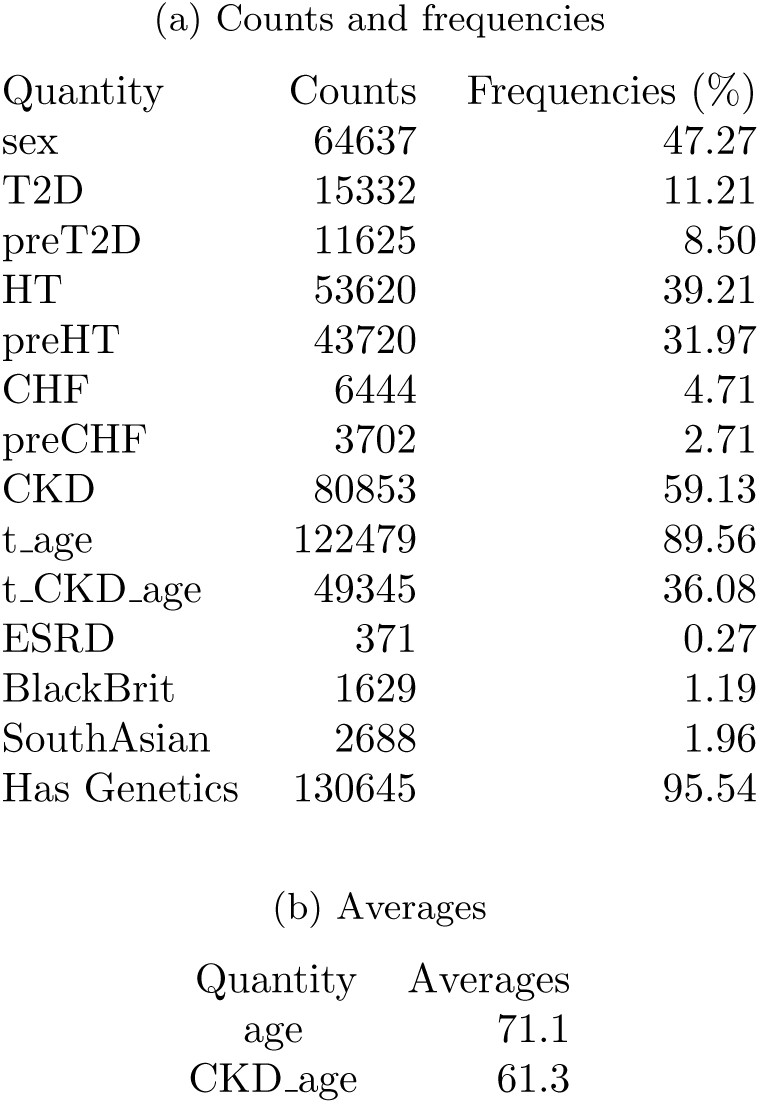
Subject Characteristics.

**Table S2:** ESRD vs. Phenotypes: Logistic Regression.

(a) CKD vs. phenotypes Logistic Regression
| Feature | OR | 95CI- | 95CI+ | P-val |
| --- | --- | --- | --- | --- |
| preHT | 2.023 | 1.972 | 2.076 | 0.000E+00 |
| preT2D | 2.112 | 2.012 | 2.218 | 2.642E-198 |
| preCHF | 1.822 | 1.675 | 1.981 | 1.026E-44 |
| t_age | 1.530 | 1.476 | 1.586 | 8.506E-119 |
| sex | 0.852 | 0.833 | 0.871 | 8.306E-45 |
| BlackBrit | 2.048 | 1.822 | 2.301 | 2.038E-33 |
| SouthAsian | 2.837 | 2.570 | 3.132 | 7.974E-95 |

| Feature | OR | 95CI- | 95CI+ | P-val |
| --- | --- | --- | --- | --- |
| preHT | 2.273 | 1.800 | 2.870 | 5.134E-12 |
| preT2D | 2.090 | 1.640 | 2.665 | 2.685E-09 |
| preCHF | 1.706 | 1.209 | 2.405 | 2.337E-03 |
| t_age | 0.734 | 0.455 | 1.184 | 2.048E-01 |
| t_CKD_age | 2.946 | 2.304 | 3.767 | 7.014E-18 |
| sex | 2.348 | 1.871 | 2.945 | 1.597E-13 |
| BlackBrit | 2.894 | 1.677 | 4.991 | 1.337E-04 |

**Table S3:** Cox regression phenotypic coefficients predicting time to ESRD.

| covariate | exp(coef) | exp(coef) lower 95% | exp(coef) upper 95% | p |
| --- | --- | --- | --- | --- |
| preHT | 0.720 | 0.444 | 1.169 | 0.184 |
| preT2D | 1.508 | 0.937 | 2.429 | 0.091 |
| preCHF | 1.760 | 0.691 | 4.480 | 0.236 |
| Thiazide | 1.134 | 0.587 | 2.188 | 0.708 |
| s_age | 0.596 | 0.408 | 0.873 | 0.008 |
| t_CKD_age | 2.301 | 1.159 | 4.570 | 0.017 |
| sex | 1.072 | 0.657 | 1.748 | 0.782 |
| BlackBrit | 0.555 | 0.103 | 2.975 | 0.492 |

**Table S4:** GO Enrichment.

| Type | Count | DE | xDE | DxE | xDxE | Pval |
| --- | --- | --- | --- | --- | --- | --- |
| Glomerulus | 25 | 19 | 6007 | 18830 | 2300270 | 1.74E-06 |
| Nephron | 40 | 14 | 6237 | 18835 | 2300040 | 1.96E-09 |
| Metanephric | 56 | 18 | 4254 | 18831 | 2302023 | 0.002693 |
| Kidney | 142 | 85 | 19964 | 18764 | 2286313 | 2.49E-11 |
| Renal | 132 | 169 | 14304 | 18680 | 2291973 | 6.07E-06 |
| Kidney/Renal | 253 | 245 | 32557 | 18604 | 2273720 | 0.203546 |

**Table S5:** ESRD vs. CKD Genotypes and Phenotypes: Logistic Regression.

(a) CKD vs. the CKD genotype and phenotypes Logistic Regression
|  | OR | 95CI- | 95CI+ | P-val |
| --- | --- | --- | --- | --- |
| preHT | 2.013 | 1.961 | 2.067 | 0.000E+00 |
| preT2D | 2.095 | 1.992 | 2.203 | 6.848E-183 |
| preCHF | 1.805 | 1.657 | 1.966 | 8.387E-42 |
| sex | 0.854 | 0.835 | 0.873 | 4.628E-42 |
| t_age | 1.561 | 1.505 | 1.621 | 2.593E-122 |
| BlackBrit | 1.395 | 1.092 | 1.783 | 7.786E-03 |
| SouthAsian | 2.712 | 2.439 | 3.017 | 1.847E-75 |
| CKDgeno | 0.898 | 0.874 | 0.922 | 1.001E-15 |

|  | OR | 95CI- | 95CI+ | P-val |
| --- | --- | --- | --- | --- |
| preHT | 2.290 | 1.801 | 2.912 | 1.403E-11 |
| preT2D | 1.978 | 1.531 | 2.556 | 1.833E-07 |
| preCHF | 1.653 | 1.148 | 2.380 | 6.914E-03 |
| t_age | 0.502 | 0.292 | 0.863 | 1.261E-02 |
| t_CKD_age | 4.313 | 3.165 | 5.878 | 2.146E-20 |
| CKDgeno | 1.239 | 0.975 | 1.575 | 7.921E-02 |
| sex | 2.423 | 1.910 | 3.073 | 2.990E-13 |
| BlackBrit | 1.456 | 0.202 | 10.491 | 7.090E-01 |
| SouthAsian | 1.221 | 0.644 | 2.314 | 5.412E-01 |

**Table S6:**
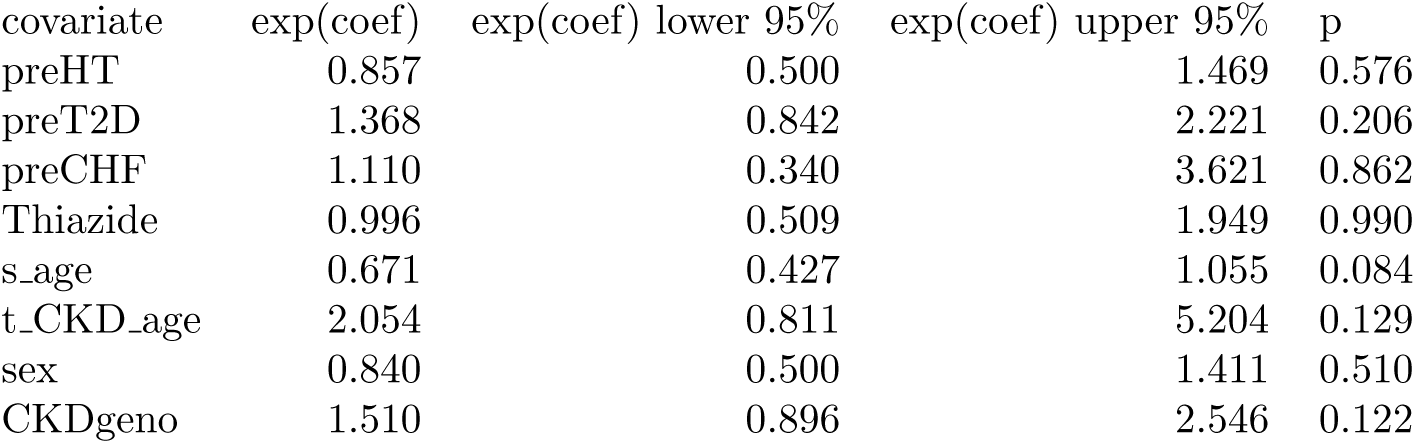
Cox regression phenotypic/genotypic coefficients predicting time to ESRD.

| covariate | exp(coef) | exp(coef) lower 95% | exp(coef) upper 95% | p |
| --- | --- | --- | --- | --- |
| preHT | 0.857 | 0.500 | 1.469 | 0.576 |
| preT2D | 1.368 | 0.842 | 2.221 | 0.206 |
| preCHF | 1.110 | 0.340 | 3.621 | 0.862 |
| Thiazide | 0.996 | 0.509 | 1.949 | 0.990 |
| s_age | 0.671 | 0.427 | 1.055 | 0.084 |
| t_CKD_age | 2.054 | 0.811 | 5.204 | 0.129 |
| sex | 0.840 | 0.500 | 1.411 | 0.510 |
| CKDgeno | 1.510 | 0.896 | 2.546 | 0.122 |

**Table S7:** GO-term SNP Counts *≥* 10 of Benjamini-Hochberg SNPs significant at 1 *×* 10^™4^.

| GO-term | Count |
| --- | --- |
| Protein binding | 104 |
| Nucleoplasm | 61 |
| Acid-thiol ligase activity | 48 |
| Fatty acid metabolic process [82] | 48 |
| Malonyl-CoA synthetase activity [83] | 48 |
| Long-chain fatty-acyl-CoA biosynthetic process [82] | 48 |
| Fatty acid biosynthetic process [82] | 48 |
| ATP binding | 48 |
| Malonate catabolic process | 48 |
| Mitochondrial matrix | 48 |
| Mitochondrion | 48 |
| Very long-chain fatty acid-CoA ligase activity [82] | 48 |
| Plasma membrane | 44 |
| Metal ion binding | 29 |
| Extracellular matrix organization [84] | 29 |
| G protein-coupled receptor activity | 23 |
| Lung connective tissue development | 22 |
| Parturition | 22 |
| Myofibroblast differentiation | 22 |
| Nipple morphogenesis | 22 |
| Hormone binding | 22 |
| Hormone-mediated signaling pathway | 22 |
| Cytosol | 21 |
| Membrane | 19 |
| miRNA-mediated gene silencing | 10 |
| Endoplasmic reticulum membrane | 10 |
| Positive regulation of nuclear-transcribed mRNA poly(A) tail shortening | 10 |
| Extracellular region | 10 |
| P-body | 10 |
| RNA binding | 10 |
| miRNA-mediated gene silencing by inhibition of translation | 10 |
| Regulation of nuclear-transcribed mRNA catabolic process, deadenylation-dependent decay | 10 |

**Table S8:** Gene SNP Counts of Benjamini-Hochberg significant SNPs at *p ≤* 1 *×* 10^™4^.

| Gene | Count |
| --- | --- |
| ACSF3 | 48 |
| RXFP1 [85] | 22 |
| TNRC6C | 10 |
| LPCAT1 [86] | 9 |
| COL4A1 [87] | 7 |
| KCNQ2 | 4 |
| LIMA1 | 4 |
| TMOD1 [88] | 2 |
| SPACA7 | 2 |
| KLF3 [89] | 2 |
| TNNT3 | 1 |
| ADGRA3 | 1 |
| DLG2 | 1 |
| AKR1C1 | 1 |
| GLI3 | 1 |
| AFM | 1 |
| GRID2IP | 1 |
| NUP205 [90] | 1 |
| KCNH5 | 1 |
| PLGRKT | 1 |
| TCP11 | 1 |
| SLC16A11 | 1 |

**Table S9:**
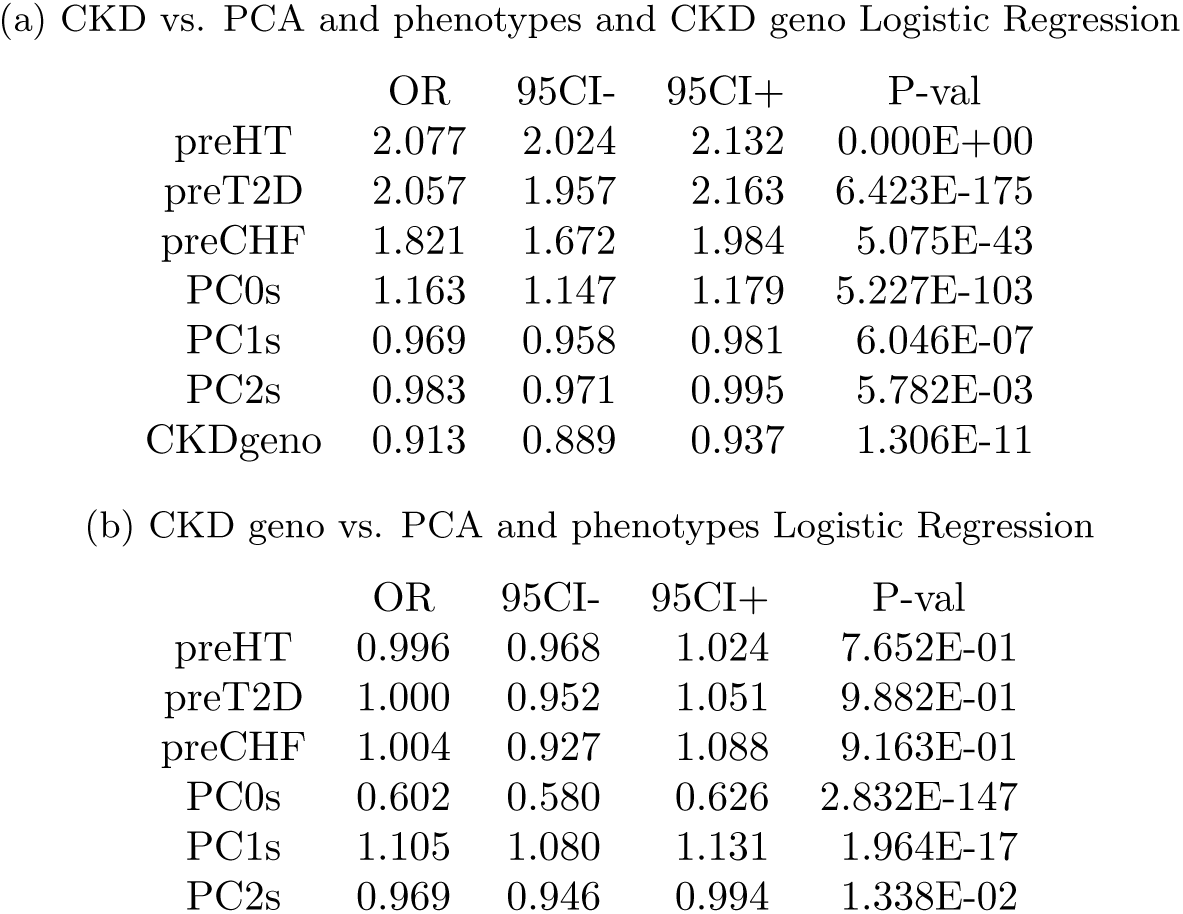
PCA impacts on CKD and relation to CKD geno.

(a) CKD vs. PCA and phenotypes and CKD geno Logistic Regression
|  | OR | 95CI- | 95CI+ | P-val |
| --- | --- | --- | --- | --- |
| preHT | 2.077 | 2.024 | 2.132 | 0.000E+00 |
| preT2D | 2.057 | 1.957 | 2.163 | 6.423E-175 |
| preCHF | 1.821 | 1.672 | 1.984 | 5.075E-43 |
| PC0s | 1.163 | 1.147 | 1.179 | 5.227E-103 |
| PC1s | 0.969 | 0.958 | 0.981 | 6.046E-07 |
| PC2s | 0.983 | 0.971 | 0.995 | 5.782E-03 |
| CKDgeno | 0.913 | 0.889 | 0.937 | 1.306E-11 |

|  | OR | 95CI- | 95CI+ | P-val |
| --- | --- | --- | --- | --- |
| preHT | 0.996 | 0.968 | 1.024 | 7.652E-01 |
| preT2D | 1.000 | 0.952 | 1.051 | 9.882E-01 |
| preCHF | 1.004 | 0.927 | 1.088 | 9.163E-01 |
| PC0s | 0.602 | 0.580 | 0.626 | 2.832E-147 |
| PC1s | 1.105 | 1.080 | 1.131 | 1.964E-17 |
| PC2s | 0.969 | 0.946 | 0.994 | 1.338E-02 |

**Table S10:** List of SNPs associated with CKD, Energy, Non-uniformity and CVD (see attached file). **ID** is the SNP identification number, **OR** is Odds Ratio, **L95** is the value for the lower bound 5% Confidence interval, **U95** is the value for the upper bound 5% Confidence interval, **P** is the raw P-value, **Pbh** is the Bonferroni corrected P-value, **symbol** is the symbol of the gene associated with the SNP, i.e in the 10kb region.

**Table S11:** Radiomics image processing table.

| Parameter | Value |
| --- | --- |
| normalizeScale | 100 |
| binWidth | 5 |
| preCrop | True |

**Table S12:** Deep learning net hyper-parameters.

| Parameter | Value |
| --- | --- |
| Learning arguments |  |
| learning rate | 1e-4 |
| weight decay | 1e-5 |
| batch size | 4 |
| Resnet arguments |  |
| first channel dim | 32 |
| first stride | 2 |
| stem kernel size | [3, 3, 3] |
| stem stride | [2, 2, 2] |
| layers | [2, 2, 2, 2] |
| Multilayer Perceptron arguments |  |
| dropout rate | 0 |
| encoder dropout rate | 0 |
| mlp hidden layers | 2 |
| mlp hidden dim | 128 |
| head hidden layers | 1 |
| head bias | True |
| use batch normalization | True |
| Vision Transformer (ViT) arguments |  |
| patch size | [16, 16, 16] |
| token dim | 512 |
| dim head | 64 |
| mlp dim | 256 |
| depth | 8 |
| heads | 8 |
| dropout | 0 |
| emb dropout | 0 |
| num cls tokens | 1 |

## Supplementary Figures

**Figure S1.**
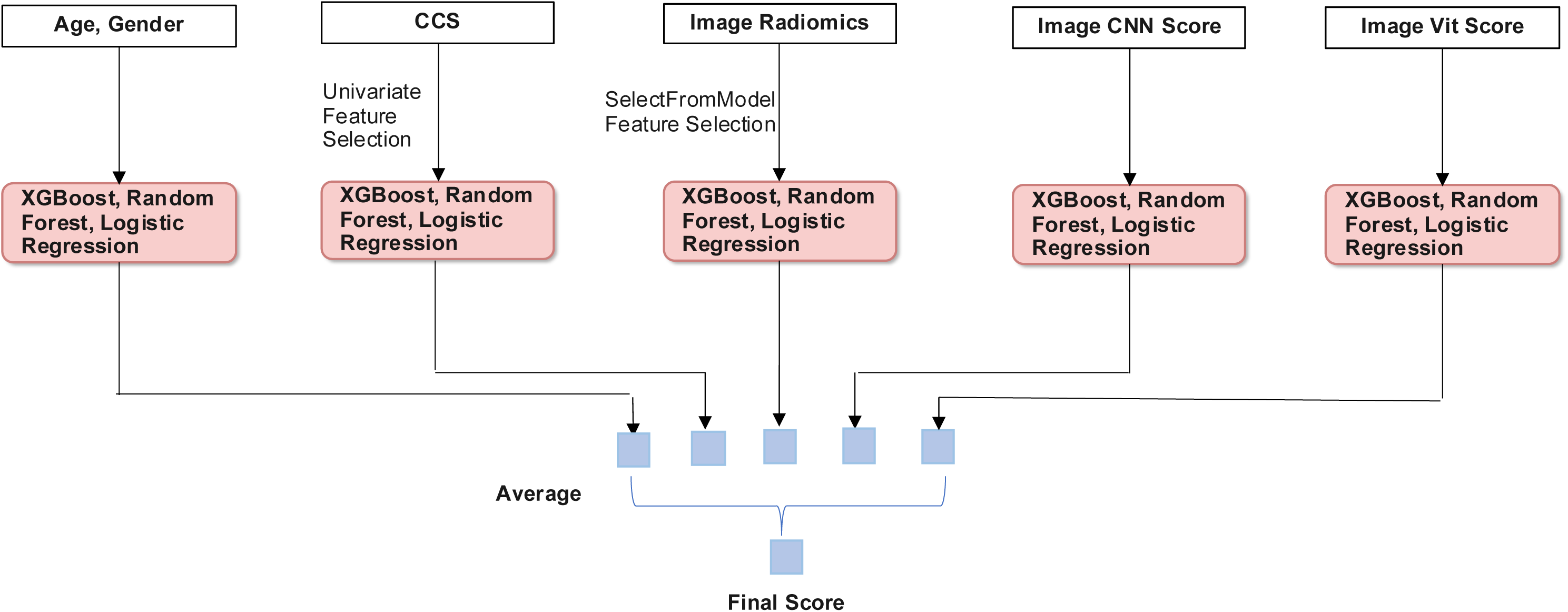
General approach for modality integration. For each of 3 data modalities and 5 models (Demographic, Clinical Codes, Image Radiomics/CNN/Vit) features were selected and implemented in a XGBoost, Random Forest, Logistic Regression classifier to predict 5-year outcome for ESRD. A voting scheme was used to determine the final prediction.

**Figure S2.**
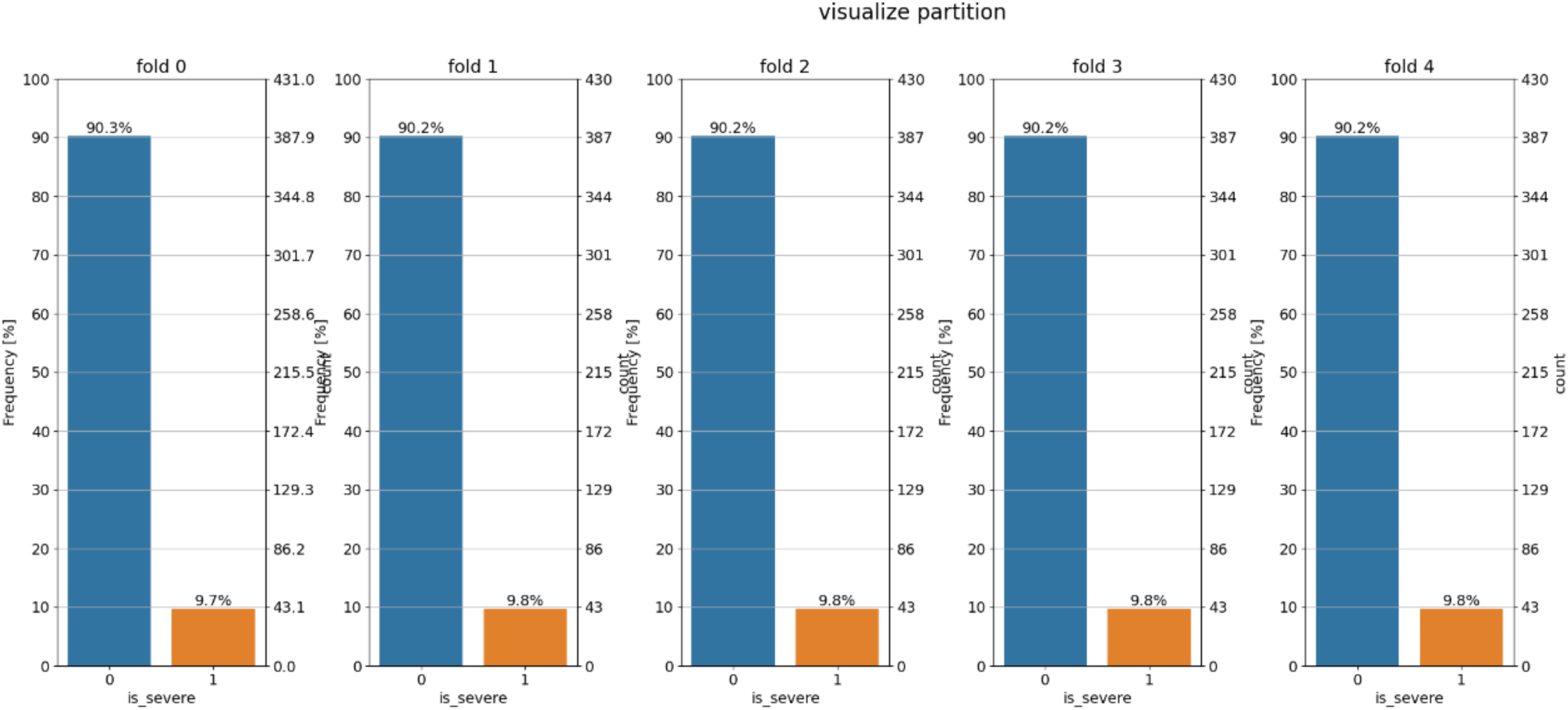
Data Splits for each data modality. Blue bars represent percentage of patients with early CKD and orange bars with ESRD for each cross-validation fold.

**Figure S3.**
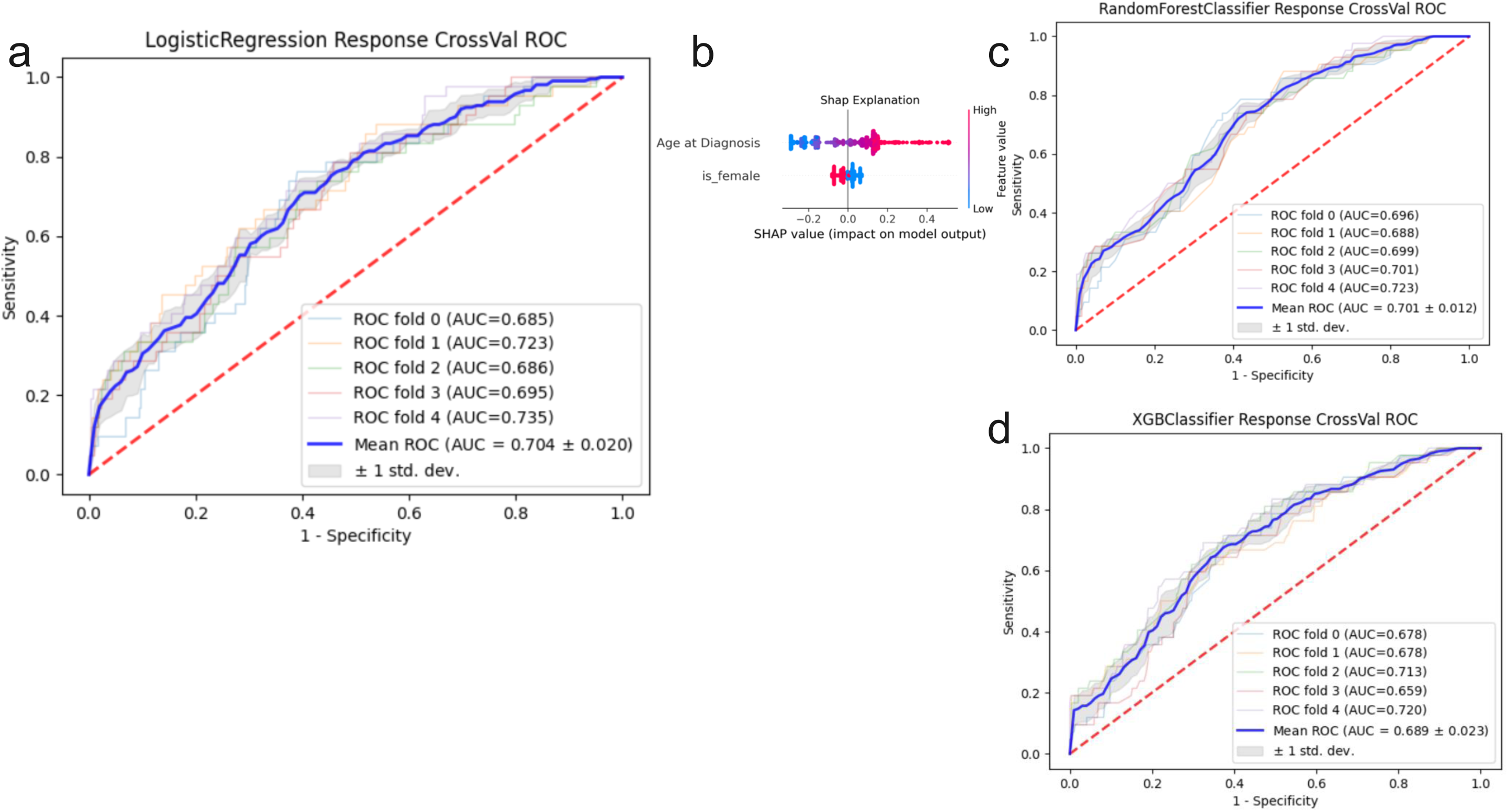
Classifiers for Demographic variables. **a.** Logistic Regression **b.** SHAP values **c.** Random Forest **d.** XGBoost classifier

**Figure S4.**
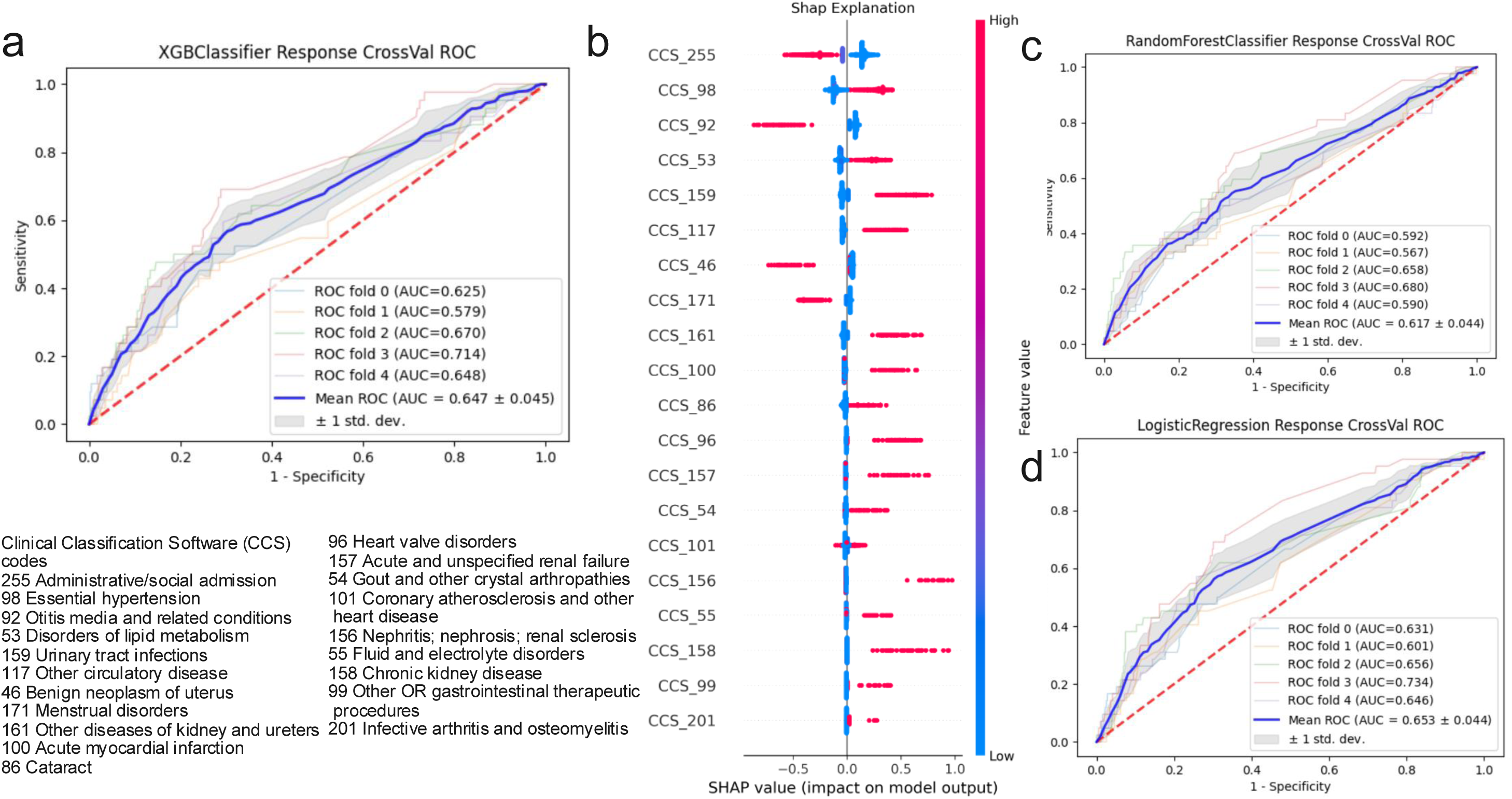
Classifiers for Clinical codes. **a.** XGBoost classifier **b.** SHAP values **c.** Random Forest **d.** Logistic Regression

**Figure S5.**
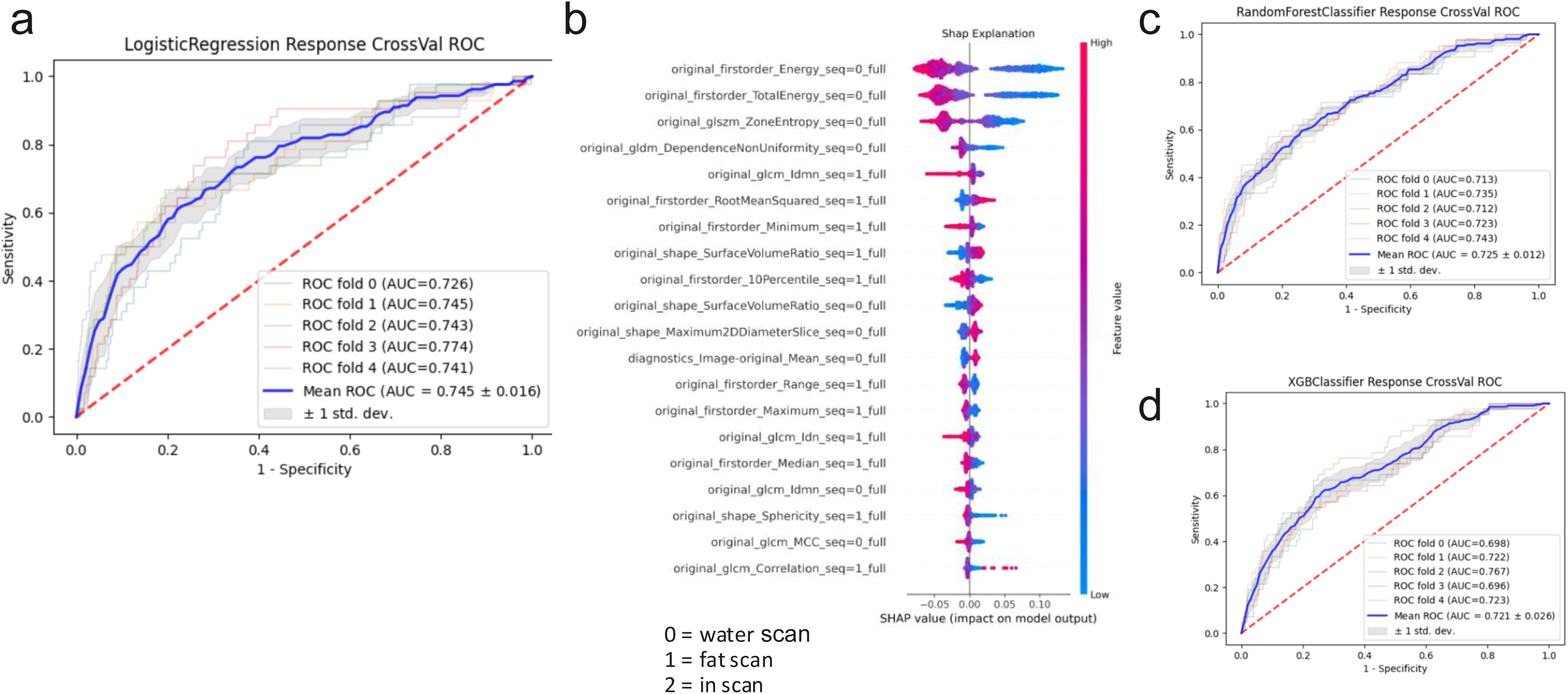
Classifiers for Image Radiomics. **a.** Logistic Regression **b.** SHAP values **c.** Random Forest **d.** XGBoost classifier

**Figure S6.**
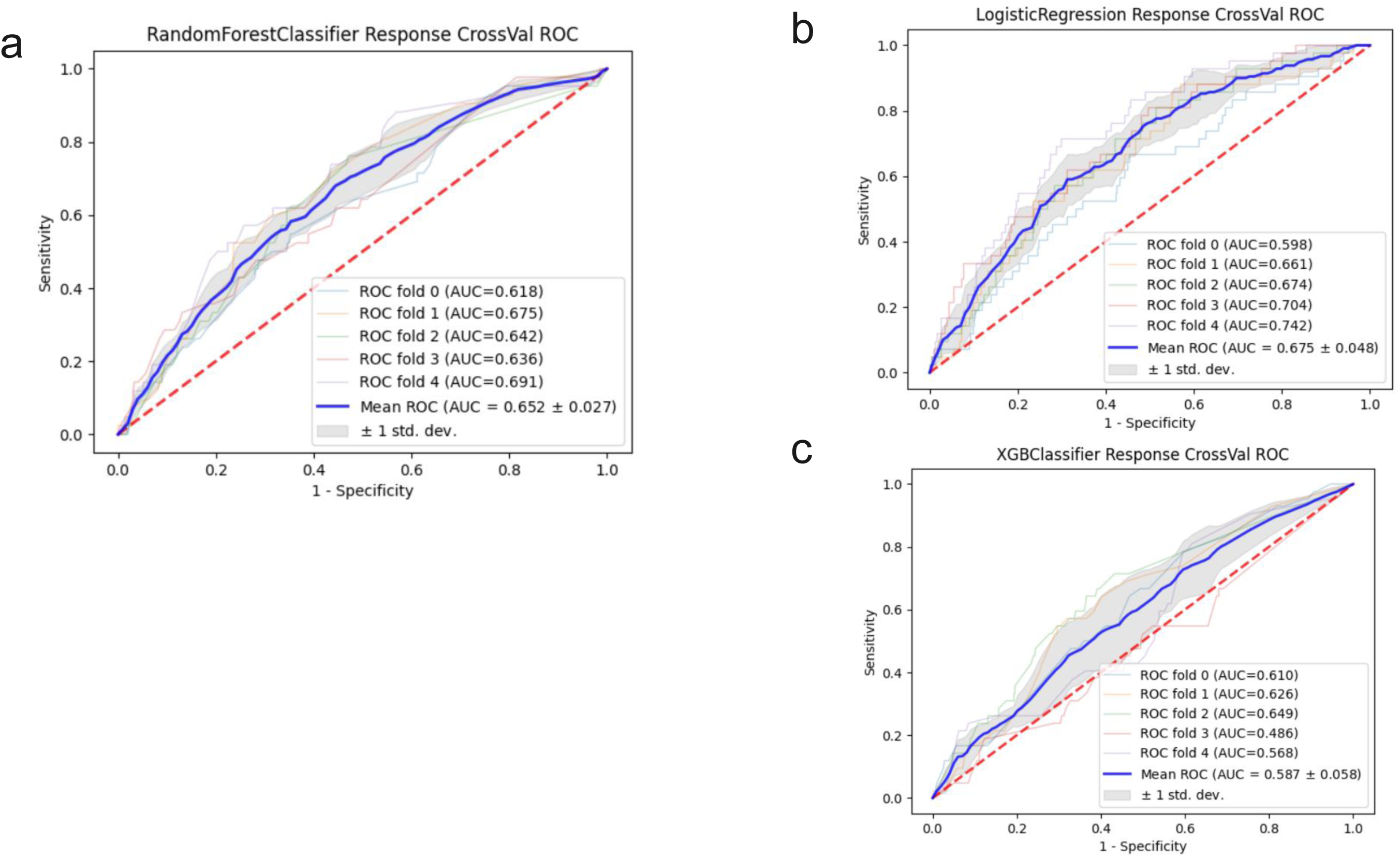
Classifiers for image CNN Score. **a.** Random Forest **b.** Logistic Regression **c.** XGBoost classifier

**Figure S7.**
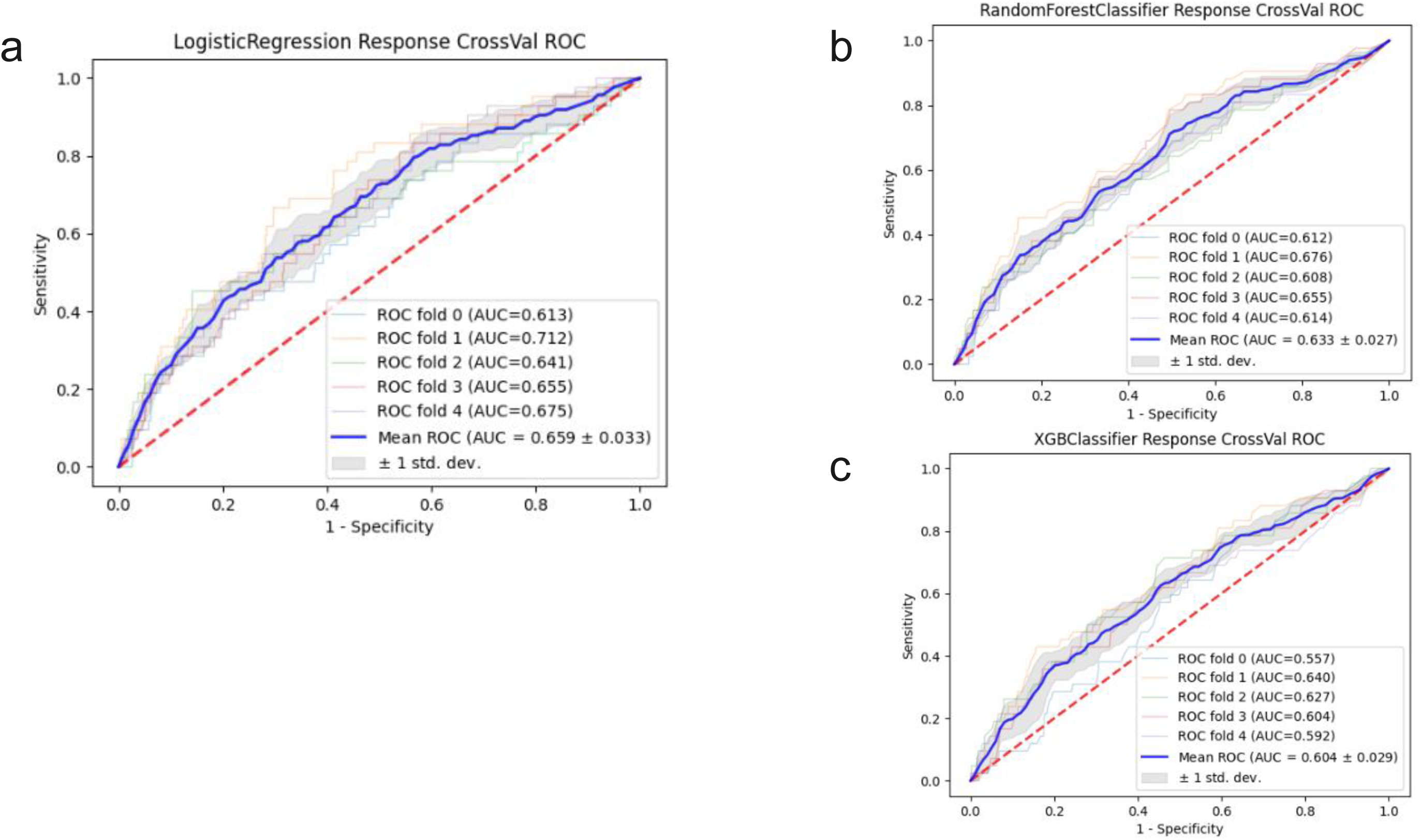
Classifiers for image ViT Score. (14/5 last epoch embeddings were used as features) **a.** Logistic Regression **b.** Random Forest **c.** XGBoost classifier

**Figure S8.**
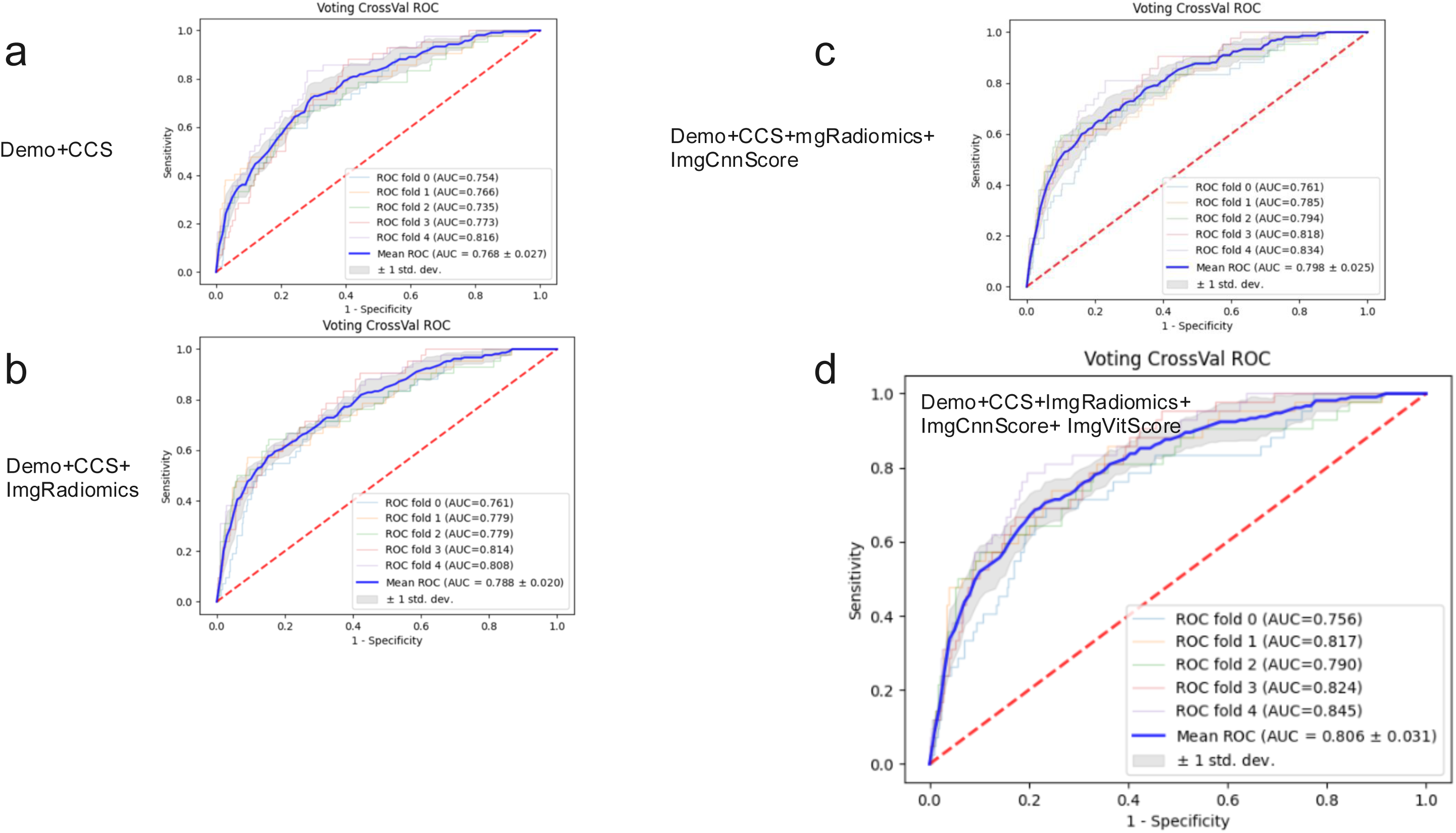
Ensemble models. Show are the results for the following combinations **a.** Demographic and CCS **b.** Demographic, CCS and Radiomics **c.** Demographic, CCS, Radiomics and CNN **d.** Demographic, CCS, Radiomics,CNN,ViT

**Fig. S9:**
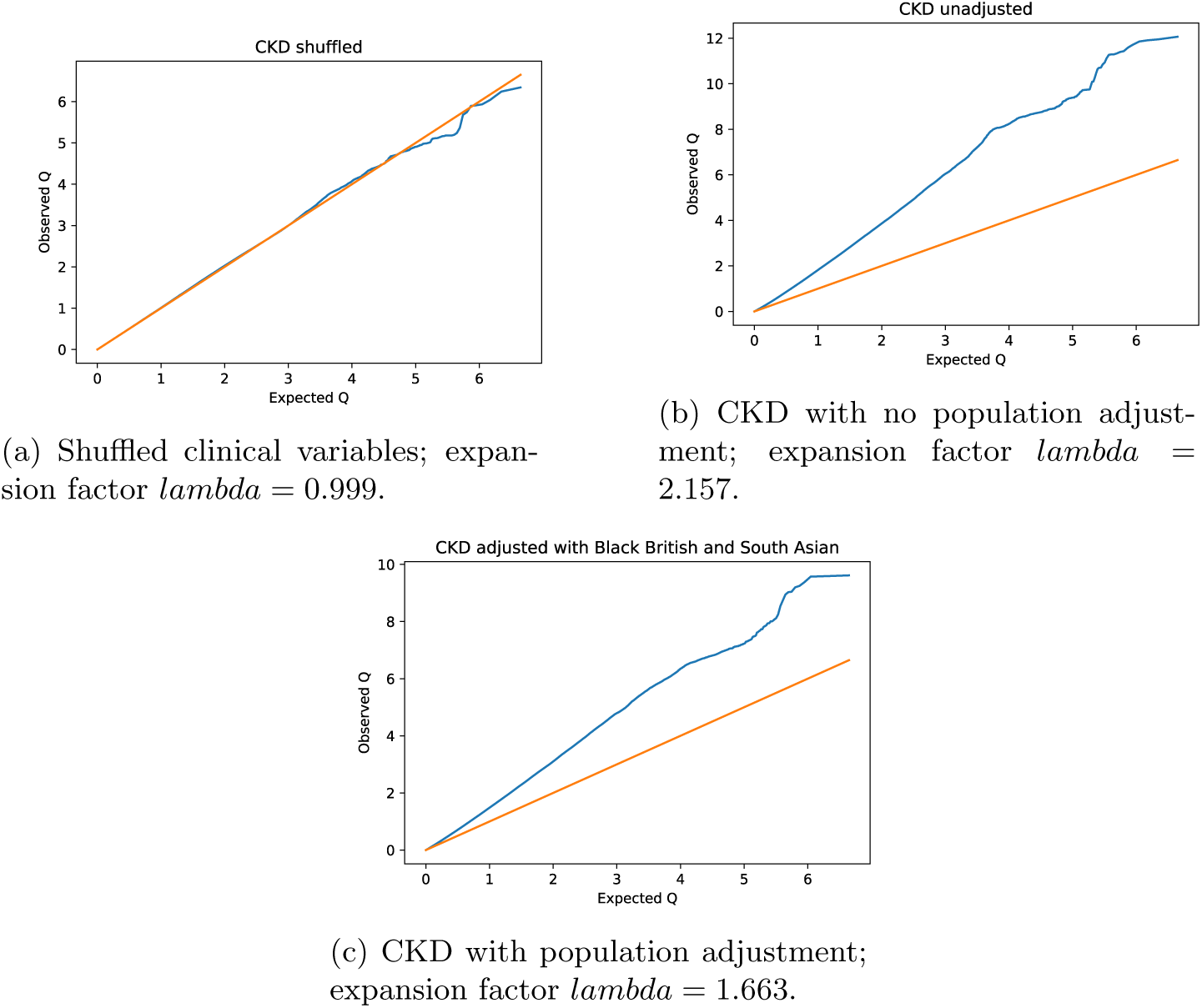
QQ plots exploring population stratification baselines, selected features, and populations.

**Fig. S10:**
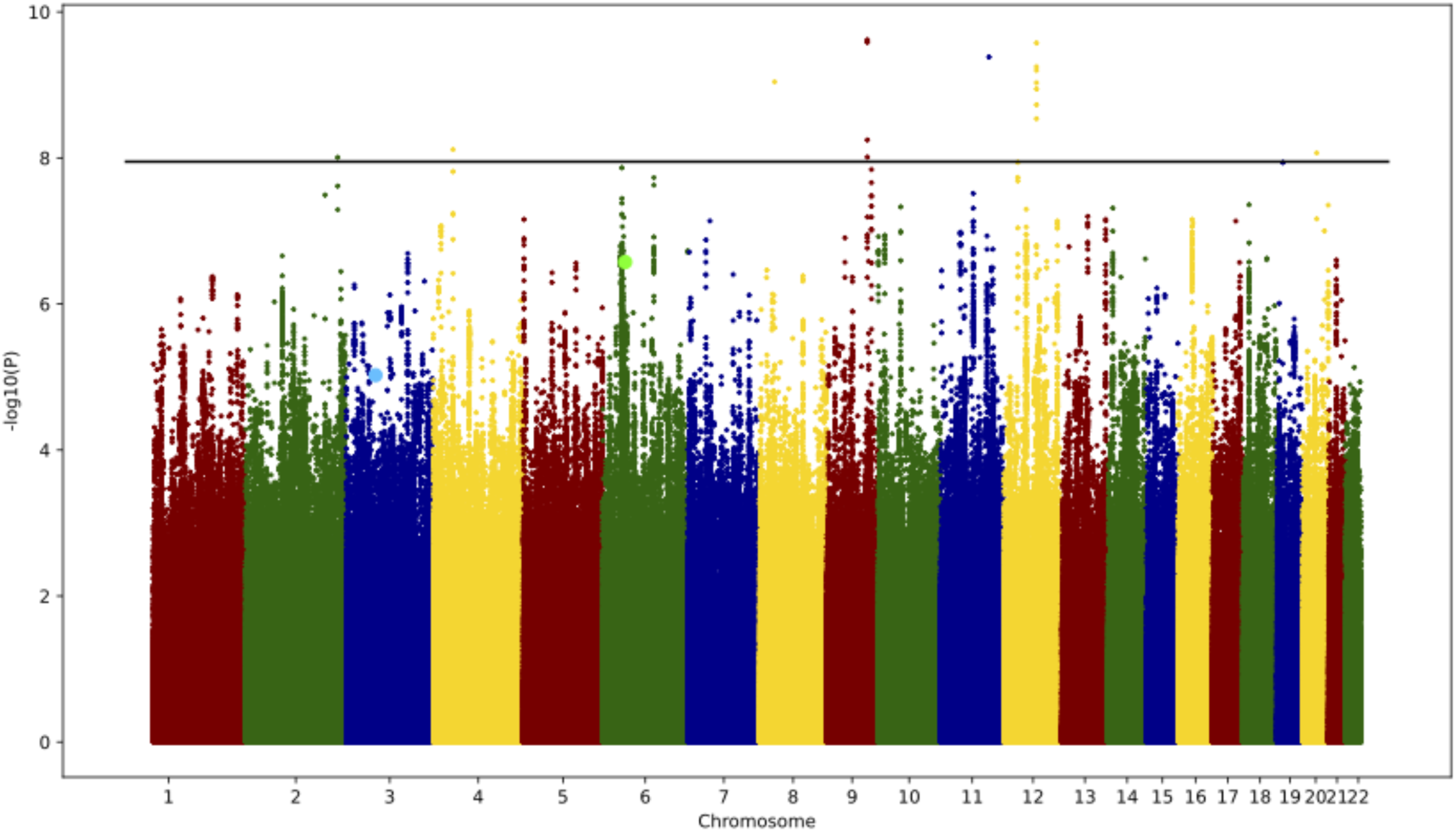
Manhattan plot for CKD logistic regression including population adjustments. The x axis is the chromosomal location of SNP and the y axis the strength of association –log10(P value). Variants rs1383063 and rs12191777 are represented by larger light blue and light green dots respectively. Line represents the limit of significance with Bonferroni correction.

**Fig. S11:**
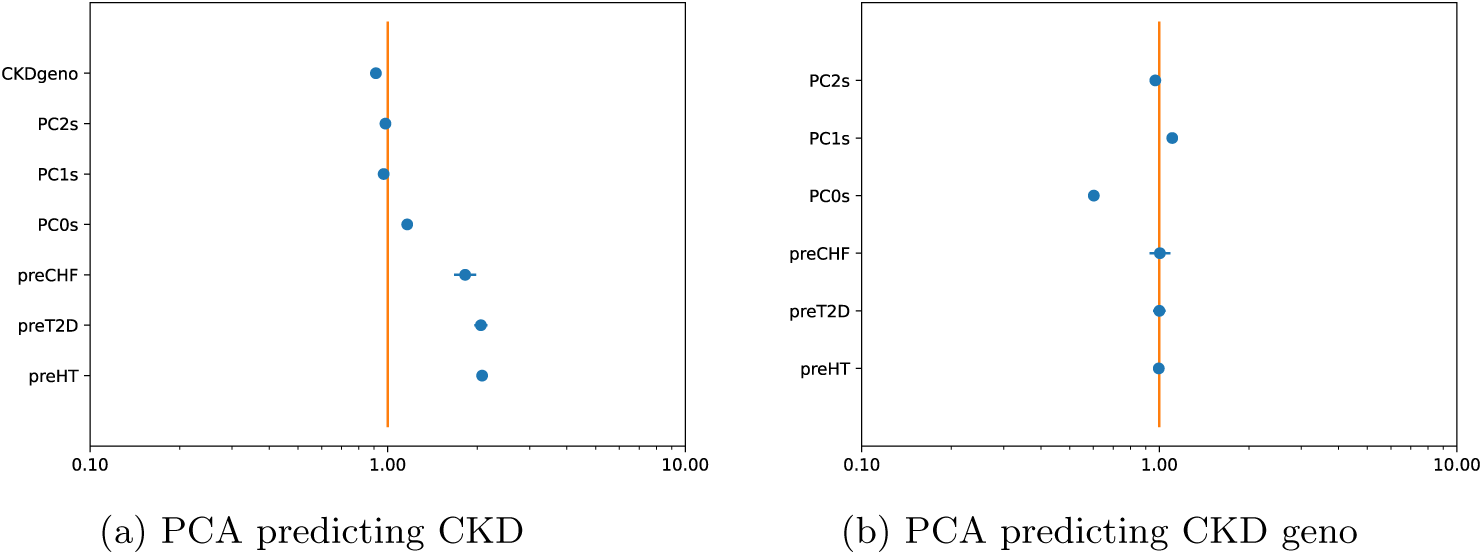
Logistic Regressions predicting CKD and CKD geno with leading principal components, pre CKD hypertension, Type II Diabetes, and Congestive Heart Failure.

**Fig. S12:**
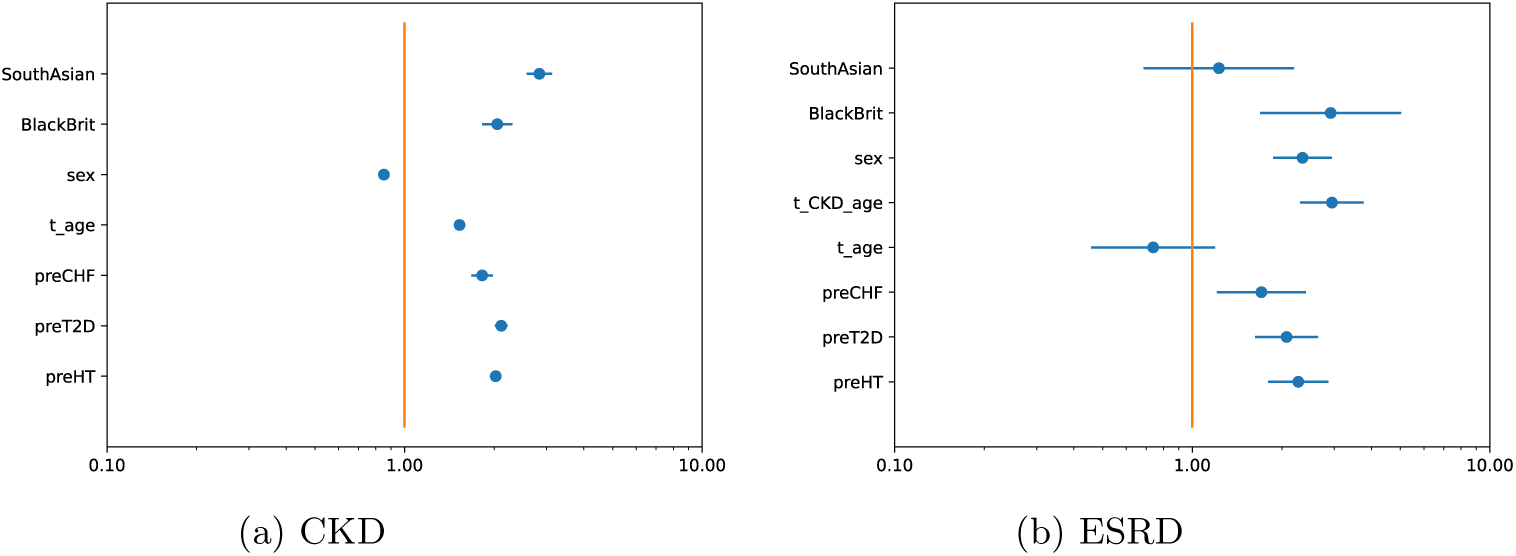
Logistic Regressions predicting CKD and ESRD with pre CKD hypertension, Type II Diabetes, Congestive Heart Failure, sex, age, and Black British status, South Asia status, and age of CKD diagnosis. t age refers to binary-threshold age of 60 years.

**Fig. S13:**
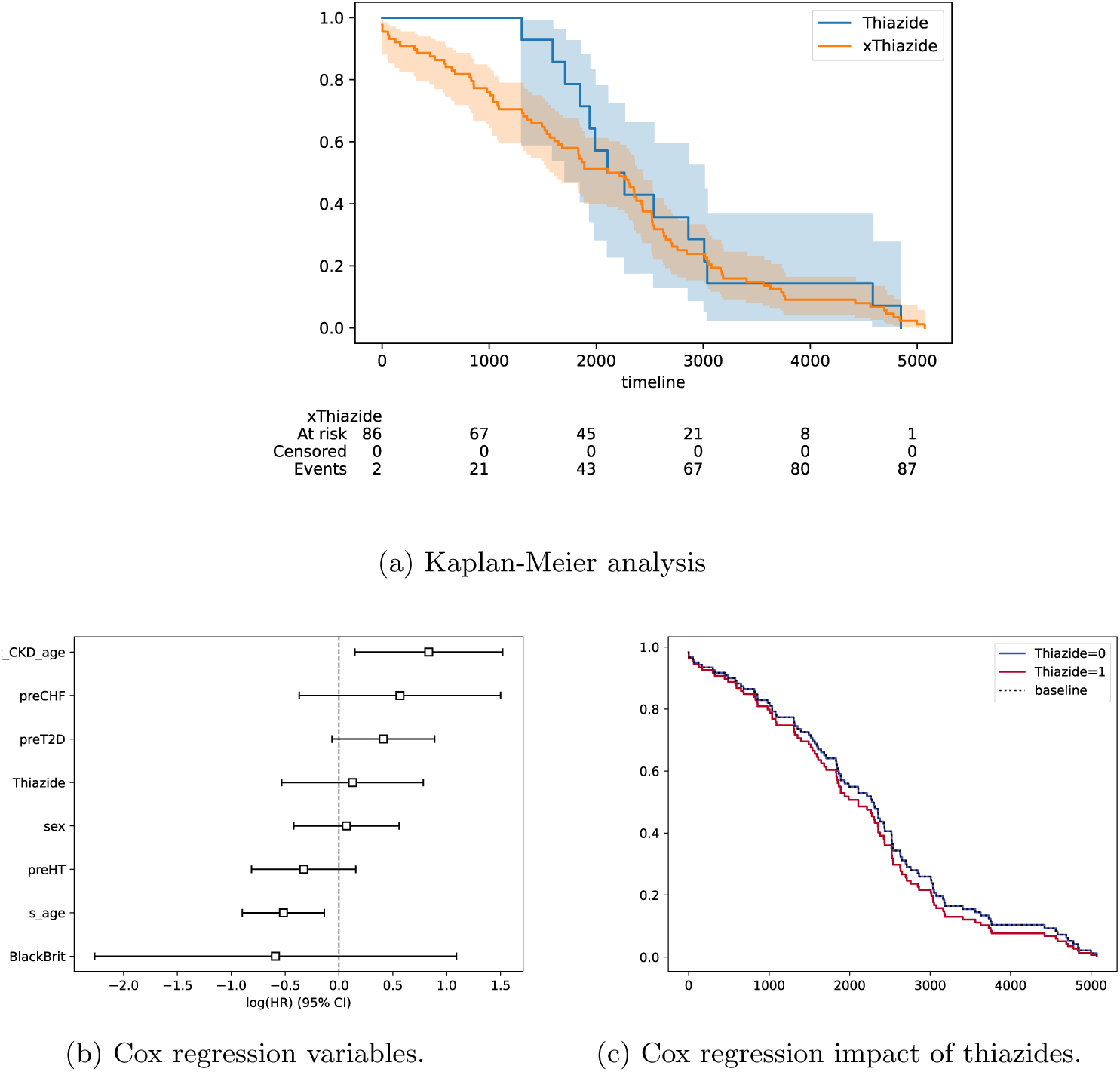
Hazard Ratio analysis of time to ESRD from CKD diagnosis. s age refers to centered and standard error scaled age.

**Fig. S14:**
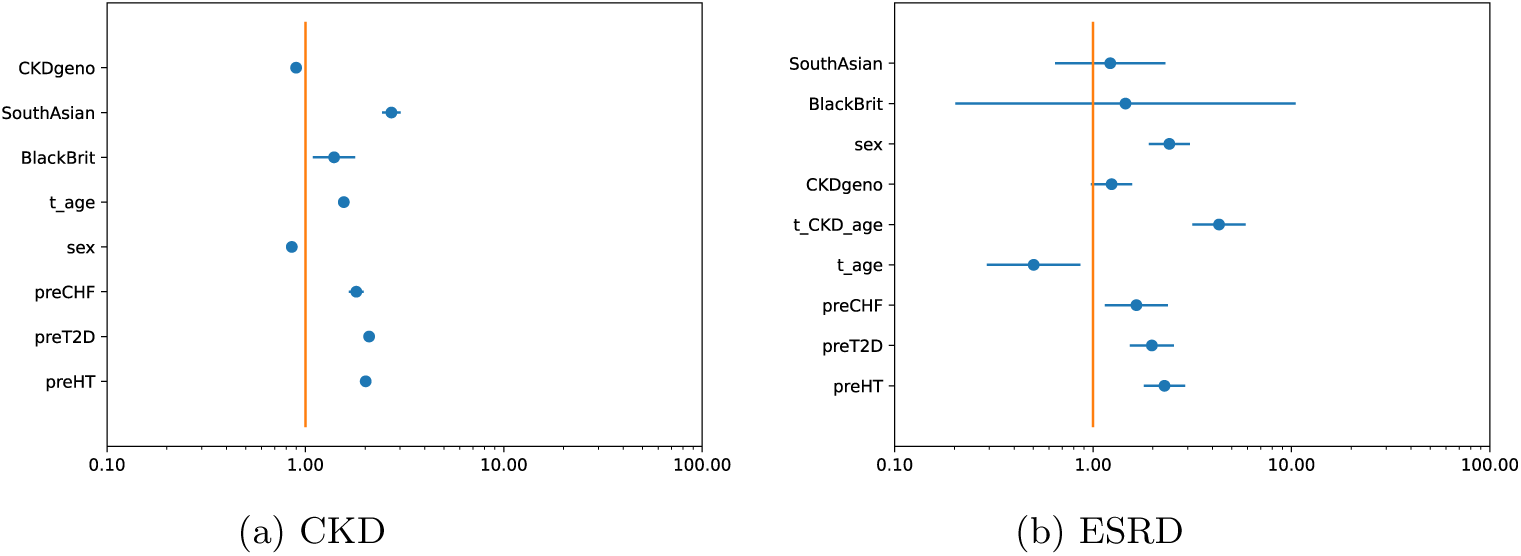
Logistic Regressions predicting CKD and ESRD with CKD significant SNPs, pre CKD hypertension, Type II Diabetes, Congestive Heart Failure, sex, age, and Black British status, South Asia status, and age of CKD diagnosis.

**Fig. S15:**
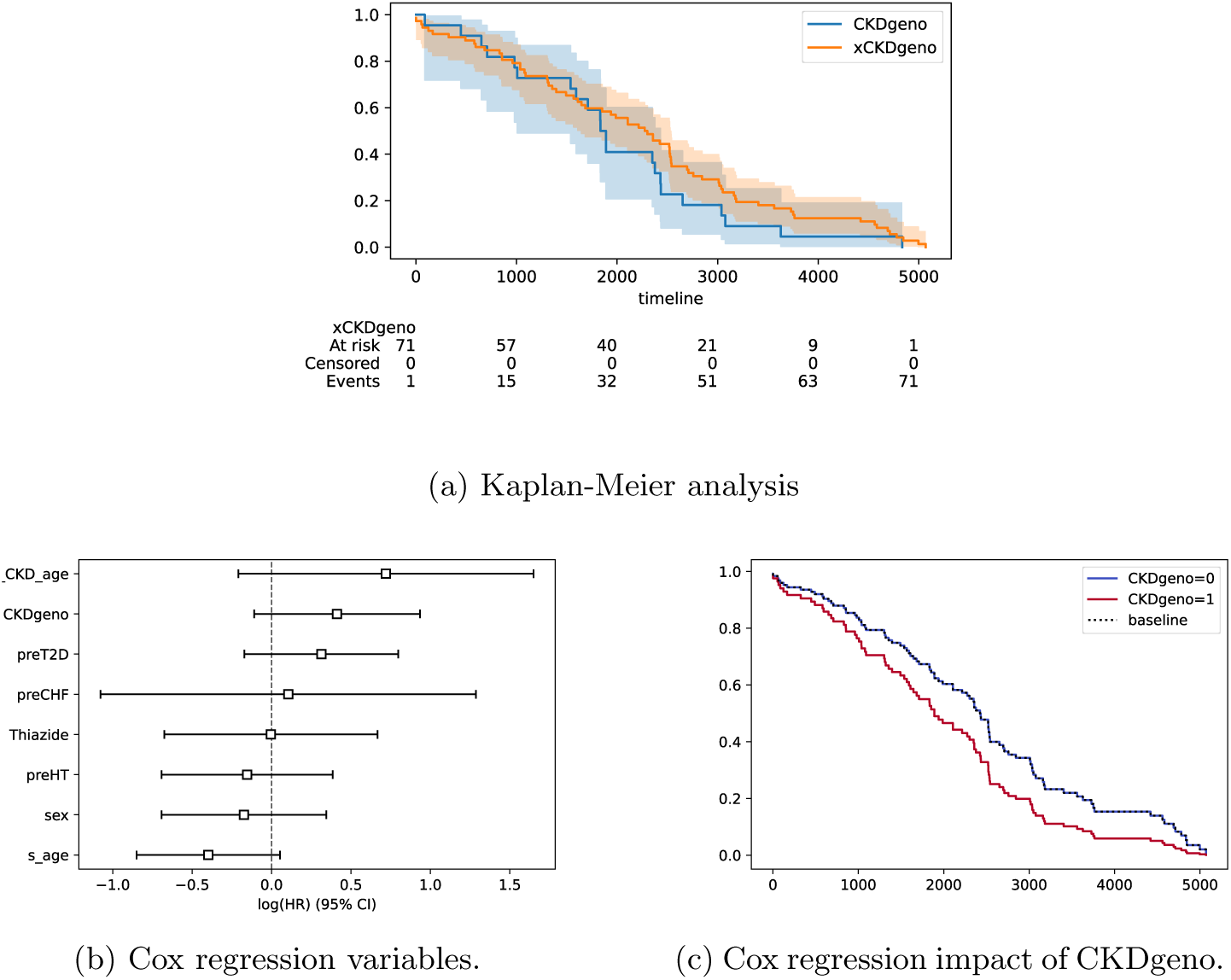
Hazard Ratio analysis of time to ESRD from CKD Genotypes. Censored or patients with ESRD happening *2:* 6000 days were excluded as also were patients were CKDGeno was not defined or missing GO-term derived kidney SNPs. 94 individuals were analyzed in total, 71 xCKDgeno and 23 CKDgeno.

**Fig. S16:**
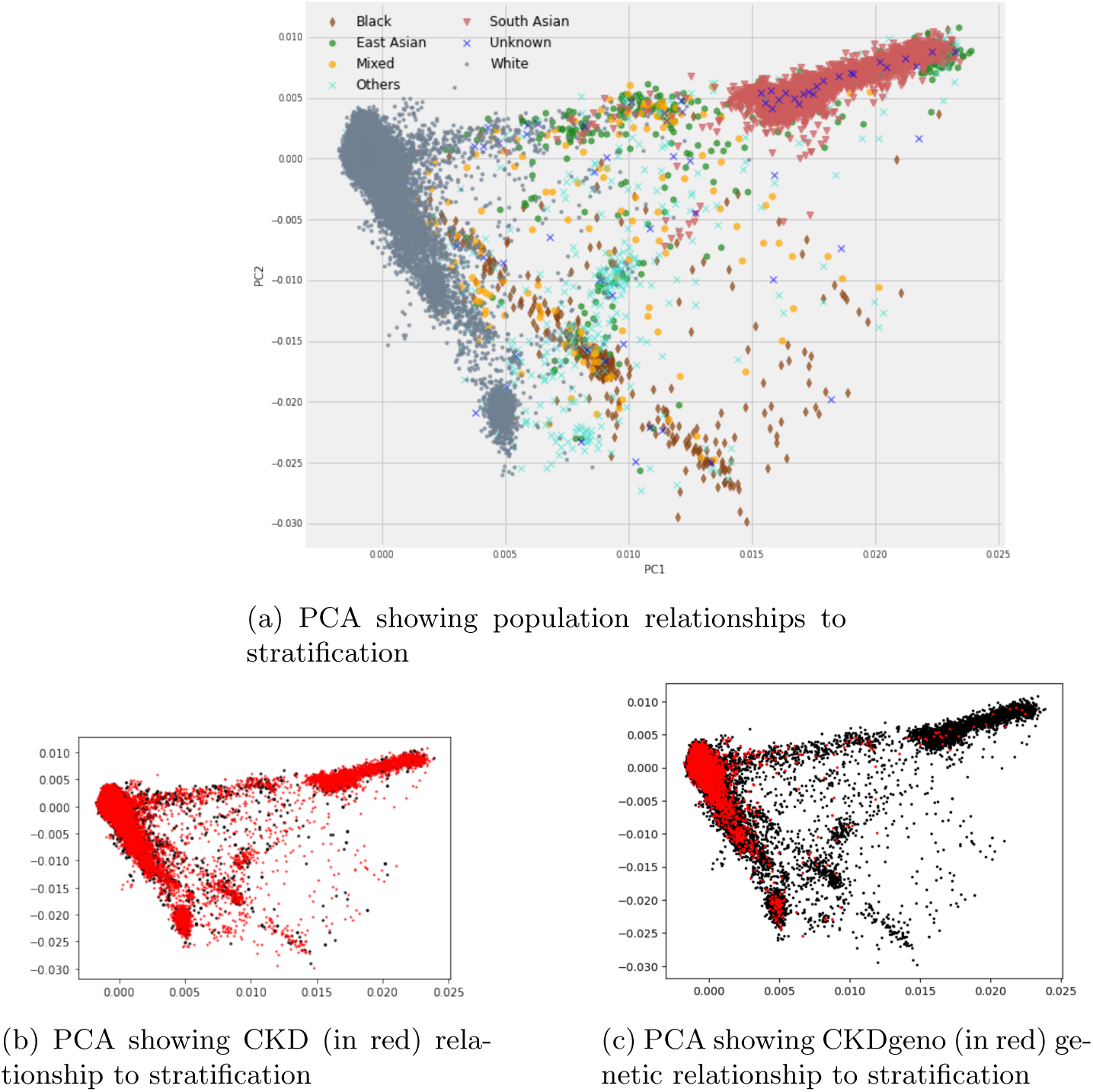
PCA showing relationships between CKD and CKDgeno to population stratification.

**Figure S17.**
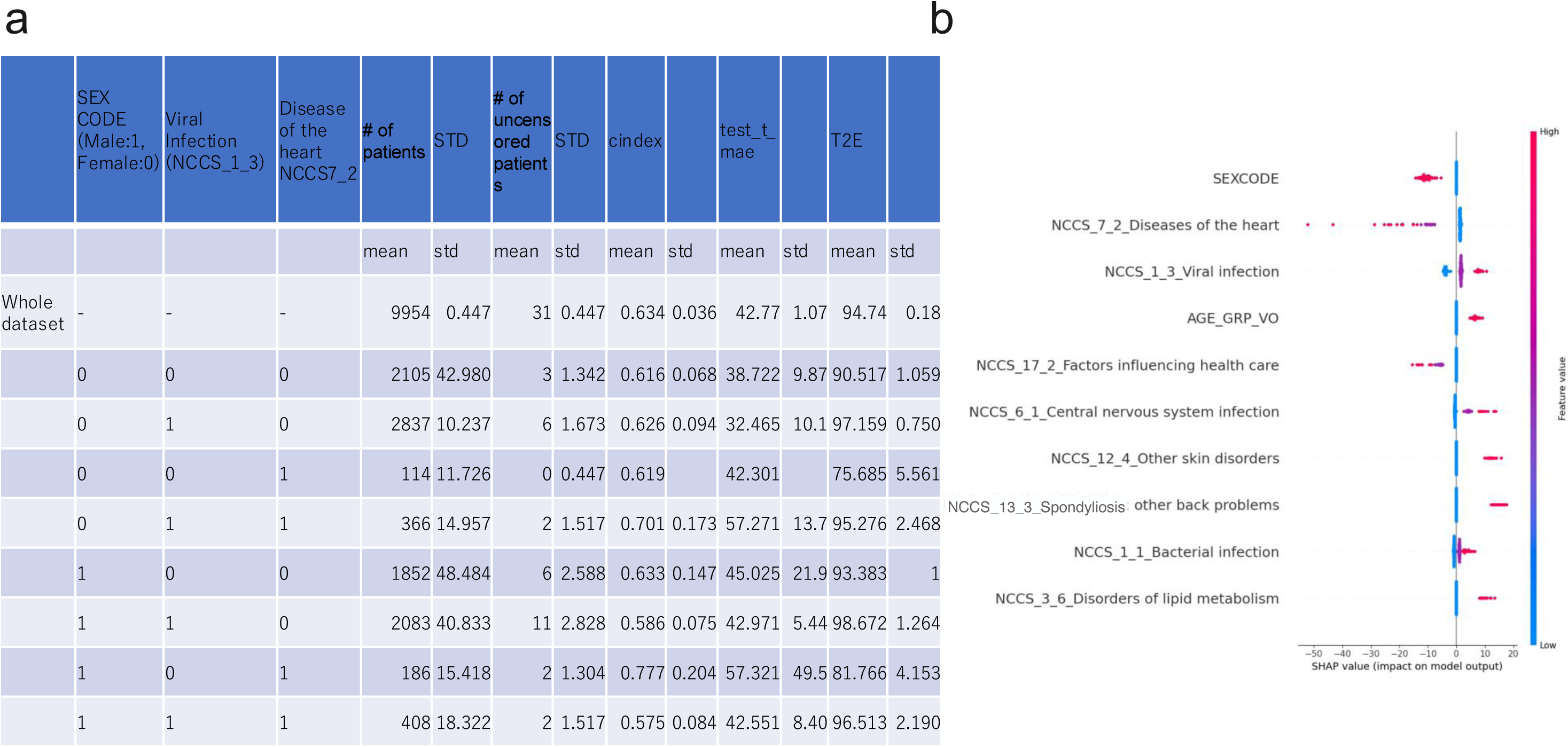
Time to ESRD predictions. **a.**CKD1&2 to ESRD prediction results of 5 fold cross-validation for test data. Only clinical data was used for the cohort of 49,744 patients with CKD. The number of patients having the top 3 features is indicated as well as the uncensored patients. Concordance Index (CI) and Mean Average Error (MAE) of prediction results are shown together with the average number of days to ESRD.**b.** SHAP results

**Figure S18.**
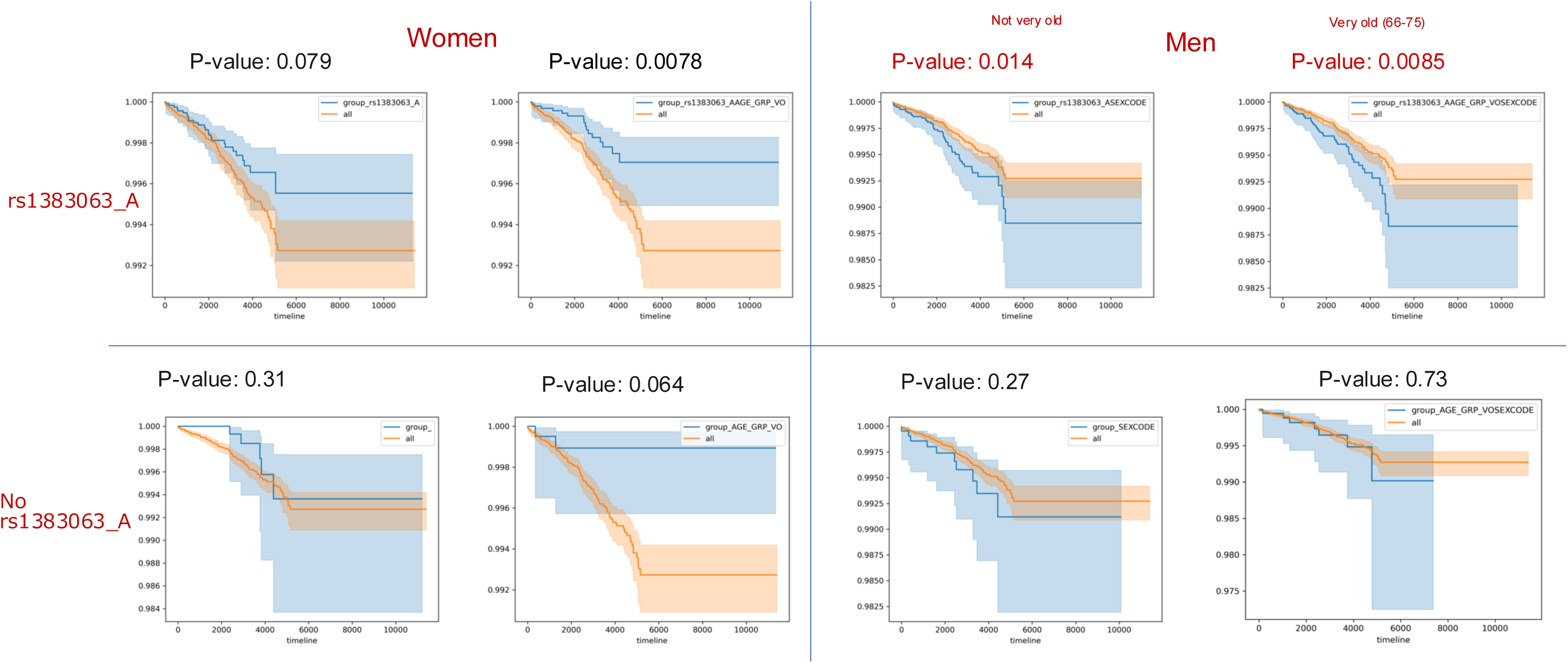
Kaplan Meier (KM) curves for survival predictions from CKD stage 1 & 2 to ESRD predictions using clinical and genomic data. Each KM curve represents parent populations in orange and subpopulations where the indicated feature is present (Sex, rs1383063_A and older age). Significant P-values are shown in red. Men with rs1383063_A have high risk of ESRD. Then, older (66-75) men with rs1383063_A have a higher risk.

**Figure S19.**
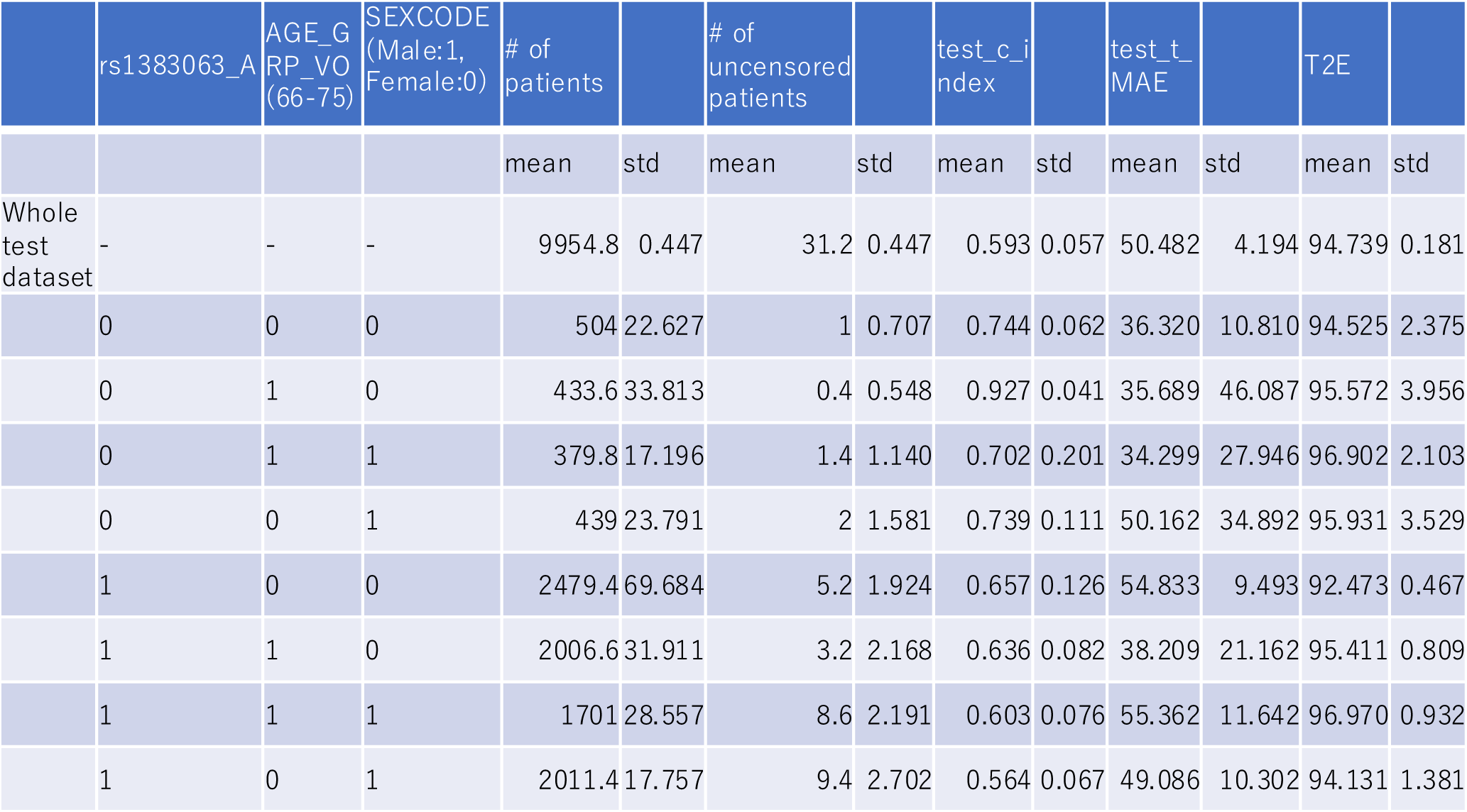
Feature information and prediction results for clinical and genomic data predictions of CKD 1&2 to ESRD time to event prediction. CKD12 results of 5cv for test data, the number of patients having the top 3 features is indicated as well as the uncensored patients. Concordance Index (CI) and Mean Average Error (MAE) of prediction results are shown together with the average number of days to ESRD.

**Figure S20.**
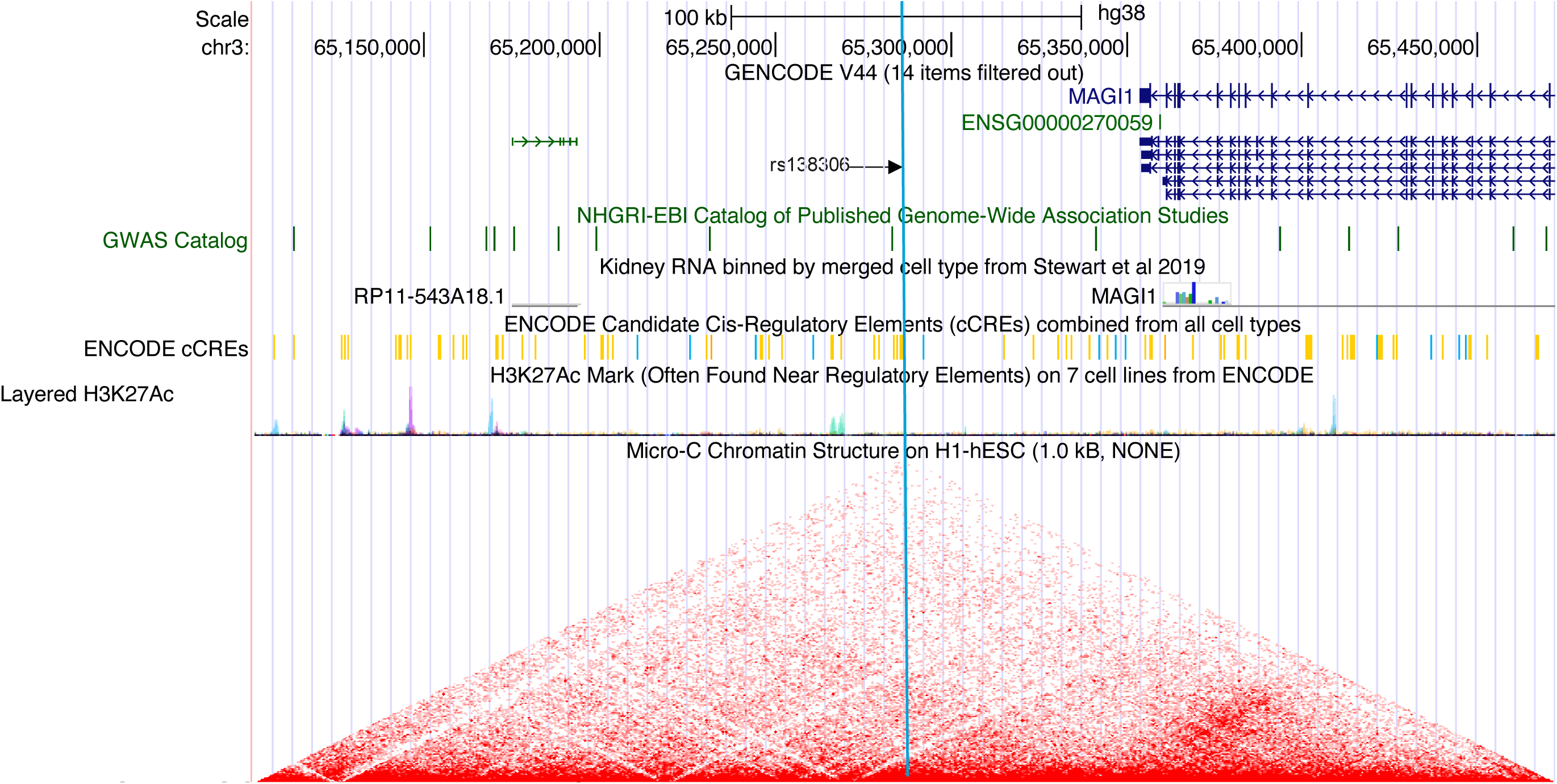
USC genome browser information on rs138306. *MAGI1* gene position is indicated in dark blue, rs138306 position is indicated by an arrow and vertical light blue line, GWAS catalog SNPs are shown in green, putative enhancers from ENCODE are shown in yellow, red triangle density indicates Topological Associated Domains as measured by microC. Scale is indicated above, as well as chromosomal coordinates.

## Footnotes

1 https://www.hcup-us.ahrq.gov/toolssoftware/ccs/AppendixCMultiDX.txt

